# Investigating the relationship between adolescent mental health and wellbeing in a nationally representative survey of 11-16 year olds in Wales

**DOI:** 10.1101/2024.12.30.24318837

**Authors:** Liam Mahedy, Basil McDonald, Giles Greene, Alisha Davies, Ann John

## Abstract

**Background:** Improving mental health (MH) and wellbeing (WB) of children is a national priority, yet there is no consensus on how these constructs are defined, nor the relationship between them. This is essential to ensure action is directed to where it has the greatest potential to prevent poorer MH and WB.

**Methods:** The School Health Research Network includes 191,975 pupils from 196 secondary schools in Wales (94% coverage of all schools). Annual pupil health survey data was used to investigate the relationship between MH (Strengths and Difficulties Questionnaire Total Difficulties (SDQTD)) and WB (Short Warwick-Edinburgh Mental Wellbeing Scale (SWEMWBS)) in a multilevel modelling framework. The primary sample size comprised any 11-to 16-year-old pupils with a pair of SWEMWBS and SDQTD scores in the 2019 or 2021 survey (N=191,975). Adjustment was made for variables hypothesised to influence MH or WB, grouped into demographic, behavioural, social, and structural domains.

**Findings:** Worsening SDQTD categories predicted lower SWEMWBS scores in a clinically significant dose-response relationship in the fully adjusted model, ranging from slightly raised SDQTD (*b* = −1.68, 95% CI = −1.76, −1.61), high SDQTD (*b* = −2.41, 95% CI = −2.51, −2.31), and very high SDQTD (*b* = −3.63, 95% CI = −3.71, −3.55). Sensitivity analysis including pupils with missing SDQTD scores showed a similar pattern of results.

**Conclusion:** These findings indicate a strong negative association between MH difficulties and WB among pupils in Wales. Determining whether treatment and prevention techniques for poor MH are shared or distinct could help allocate resources and efforts efficiently.

## Introduction

Understanding the relationship between mental health (MH) and mental wellbeing (WB) in children and young people is crucial to inform policy development to prevent poor MH, promote good MH and to support and care for those in need across health, social care and other sectors including education (Ford, John, & Gunnell, 2021; Weare, 2017). Supporting WB is thought to contribute to protecting MH, suggesting that interventions aimed at enhancing WB could have protective effects against MH disorders (Patalay & Fitzsimons, 2016a). Despite the acknowledged importance of both constructs for children and young people (World Health Organization, 2021), there is limited knowledge on how MH and WB are defined, and what elements of overlap or distinction there are between the two.

Issues arise from the lack of consensus on the definitions of MH and WB, including whether they represent opposing ends of a single continuum and how they can be distinctly differentiated from one another. Evidence from child and adolescent studies demonstrated shared variance between MH difficulties and WB (Kendler, Myers, & Keyes, 2011; Lereya, Patalay, & Deighton, 2022), albeit sometimes relatively minimal (Patalay & Fitzsimons, 2016b). Others prefer a dual continua model which views MH difficulties and WB as two separate concepts (Suldo & Shaffer, 2008; Westerhof & Keyes, 2010). The extent to which these constructs may overlap and interact is unclear as evidenced by inconsistent findings. For instance, studies have suggested individuals could experience good WB despite suffering MH difficulties (Kinderman et al., 2015; Sharpe et al., 2016), while others suggested the opposite, in that individuals could enjoy good MH despite experiencing poor WB (Keyes, 2005).

Methodological concerns in existing studies create additional complexity in understanding the relationship between MH and WB. For example, the Millennium Cohort Study (Connelly & Platt, 2014) used in (Patalay & Fitzsimons, 2016b), measures child WB as reported by children, but child MH as reported by parents, potentially leading to inter-observer bias or measurement bias. Data from the HeadStart programme (Big Lottery Fund, 2016), used in(Lereya et al., 2022) did not use a representative national sample as the programme focussed specifically on disadvantaged areas. Another difficulty is that many of the studies above have not consistently controlled for all relevant domains (e.g., demographic, behavioural, and social) at the same time potentially introducing bias due to confounding, and limiting the accuracy of their findings (Patton et al., 2016).

The aim of this study was to develop our understanding of the relationship between MH and WB by using pupil reported data to establish the relationship between these two constructs. This study will seek to overcome limitations outlined above by using validated measures in the pupil reported data of the School Health Research Network (SHRN), a Wales whole population based survey of schoolchildren aged 11-16 years, responding to questions about their MH and WB, and including comprehensive data on social and economic confounders at an individual, family and household level. This approach enhances the generalisability of the findings and provides robust evidence to inform future research and policy decisions. The main relevant previous study using this data (Lowthian et al., 2021) examined student WB by socioeconomic status (measured using family affluence, free school meal eligibility and parental employment), but did not control for non-economic factors, such as family and peer support or bullying.

## METHOD

### Data

This cross-sectional study is a secondary data analysis of two iterations of the SHRN biennial Student Health and Wellbeing survey, 2019/20 (Hammond, Hallingberg, & Moore, 2020), and 2021/22 (Student Health and Wellbeing in Wales, 2022). SHRN was established in 2013 by Cardiff University, Welsh Government, Public Health Wales, and other partners with the intention of understanding and improving the health and wellbeing of children and adolescents in Wales through supporting evidence-based practice and policy making. Responses could be linked at individual-level across survey iterations. SHRN is an electronic, closed-response, self-completion survey encompassing a wide range of questions on adolescent health and risk behaviours.

### Design and Sample

#### Design

This study utilised cross-sectional data from the SHRN in Wales, collected at two timepoints: Wave 1 collected from September 2019 to January 2020, and Wave 2 collected from September 2021 to January 2022. SHRN included Strengths and Difficulties Questionnaire (SDQ) (Robert Goodman, 1997) and mental wellbeing (assessed using the Short Warwick-Edinburgh Mental Wellbeing Scale (SWEMWBS) (Stewart-Brown et al., 2009). The SDQ was chosen as the exposure and SWEMWBS as the outcome, based on the theoretical relationship between the two constructs (de Cates, Stranges, Blake, & Weich, 2015).

##### Exposure

###### Strengths and Difficulties Questionnaire (SDQ)

The SDQ is a short behavioural questionnaire for 2-17-year-olds, commonly used as a screening tool for MH problems. It has been shown to be effective in differentiating cases with mental illness from those without mental illness (AUC 0.83) and in identifying MH conditions, such as oppositional or conduct disorders (AUC 0.77) (Becker, Hagenberg, Roessner, Woerner, & Rothenberger, 2004). SDQ Total Difficulties (SDQTD) was calculated by combining four subscales: (i) emotional problems, (ii) conduct problems, (iii) hyperactivity, (iv) and peer problems. Continuous scores assessing the SDQTD score were previously made into categorical variables, each with four categories for each of the five subscales: ‘close to average’, ‘slightly raised’, ‘high’ and ‘very high’ according to the usual scoring criteria (Goodman, 2001). Further information is included in Supporting Information (Exposure section).

##### Outcome

###### Short Warwick-Edinburgh Mental Wellbeing Scale (SWEMWBS)

The Short Warwick-Edinburgh Mental Wellbeing Scale (SWEMWBS) was designed to monitor WB in the general population. It is an abbreviated, 7-item version of the original 14-item Warwick-Edinburgh Mental Wellbeing Scale (WEMWBS). The SWEMWBS is a continuous measure with scores range from 7, indicating probable clinical depression, to 35, indicating high WB. The minimum statistically important change detected by SWEMWBS has variously been calculated as 1 or 3 (Shah, Cader, Andrews, Wijesekera, & Stewart-Brown, 2018). SWEWBS was used a continuous measure to retain information, allowing for a detailed analysis that captures the full range of variability within the measure. To improve clinical understanding of the SWEMWBS scores, findings were interpreted in relation to previously identified classifications (Ng Fat, Scholes, Boniface, Mindell, & Stewart-Brown, 2017).

#### Sample

This study used data from the 2019/20 and 2021/22 surveys. In the 2019/20 survey, 119,388 pupils from 198 out of 210 schools in years 7 to 11 responded to the survey (77% response rate). In the 2021/22 survey, 123,204 pupils from 202/212 schools in years 7 to 11 responded to the survey (75% response rate). Students who completed either the 2019/20 or 2021/22 surveys were included in the study. On the occasion where students completed both surveys, their responses from the 2021/22 survey were included in the analysis.

##### Covariates

Demographic, behavioural and social variables associated with children and adolescents’ MH and WB were included as covariates based on existing evidence (Kirkbride et al., 2024; Stewart-Brown, Samaraweera, Taggart, Kandala, & Stranges, 2015). The ‘demographic’ domain included age, sex, and year of assessment. The ‘behavioural’ domain included tobacco smoking, alcohol consumption, drug use, physical activity, sleep difficulties, and body image. The ‘social’ domain contained family practical support, support from friends, children’s living arrangements, having been bullied, school support, care from teachers, and the Family Affluence Scale (Currie et al., 2008). Further information is presented in the Supporting Information (Covariates section). To assess the impact of each of the domains, covariates were included in a hierarchical regression approach i) unadjusted model (model 1); ii) demographic domain (model 2); behavioural domain (model 3); and social domain (model 4).

#### Analysis

##### Primary analysis

A series of univariable and multivariable multilevel models were conducted to examine the association between MH (assessed using the SDQ) and WB (assessed using the SWEMWBS) while accounting for clustering of individuals within schools. Clustering by school was included in the regression models to account for the greater similarities between students at the same school compared to students at differing schools. Students within each school share common experiences, such as the education environment, teaching practices, and socio-economic factors. Fixed effects were estimated to examine the average effect of MH difficulties (i.e., SDQ) on WB (i.e., SWEMWBS). Unstandardised coefficients with 95% confidence intervals were used and can be interpreted as a change in the different exposure categories compared to the reference category being associated with a one-point change in the outcome. Random effects account for variability in the outcome across different schools. Results using multiple imputed estimates are reported as the primary results (N=191,975). Analyses were conducted using R version 4.4.0 (R Core Team, 2024).

##### Secondary analysis

Analysing the SDQ subscales as exposures, allows for a more detailed understanding of specific areas of MH difficulties, which will help to identify differential impacts on wellbeing. The same approach to including covariates in a hierarchy was conducted as used in the primary analysis. To ensure the robustness of our findings, Bonferroni correction was applied to account for multiple comparisons. Given that separate multilevel regression models were conducted for each of the SDQ subscales, significance threshold were adjusted by dividing the conventional alpha level (0.05) by the number of tests performed (i.e., 5), reducing the risk of Type I errors. Results using multiple imputation estimates were reported as the secondary analyses (N=191,975).

##### Dealing with missing data

To maximise sample size, this analysis was based on core questions asked to all individuals. Since using complete analysis can result in biased estimates (Sterne et al., 2009), we examined possible effects of missing data. The missing at random (MAR) assumption was made more plausible by the inclusion of socio-demographic variables in the fully adjusted model. Multiple imputation (MI) (Little & Rubin, 2002) was used to handle missing data. Supporting Information Table S1 shows evidence of a relationship between variables from all the demographic, behavioural, and social domains and missing data on the SWEMWBS measure, suggesting data fulfilled the MAR assumption. MI was based on 191,975 individuals who provided complete information on SWEMWBS at either of the 2019/20 or 2021/22 surveys and who had incomplete information on the exposure and covariates. The imputation model contained the SWEMWBS, SDQTD score, SDQ subscales, and all covariates. The imputation process generated 25 datasets by 5 cycles of regression models.

##### Sensitivity analysis

Sensitivity analyses were conducted to ensure the robustness of the findings. First, the analyses outlined above were conducted on complete cases (N=114,439). Second, analyses were conducted on an MI sample that consisted of individuals who had incomplete information on SWEMWBS, SDQTD, and covariates (N=212,975). Third, analyses were conducted on 2019/20 year of assessment for individuals who contained complete information on the SWEMWBS and partial information on the exposure and covariates (identical to the criteria for the primary analyses). This approach was adopted to ensure consistency between the pre- and post-Covid years. Fourth, analyses were conducted on 2021/22 year of assessment for individuals who contained complete information on the SWEMWBS and partial information on the exposure and covariates (identical to the criteria for the primary analyses).

##### Ethics

Details regarding the ethical considerations and approvals for this study are available here (Page et al., 2024).

## RESULTS

The sample consisted of 191,975 individuals from 196 schools (Males: N=94,810 (49.4%), Females: N=92,987 (48.4%), and Non-binary: N=4,178 (2.2%)). Age was categorised into two-year groups (11-12 years N=69,278 (36.1%), 13-14 years: N=78,503 (40.9%), and 15-16 years: N=44,194 (23.0%)). The SWEMWBS ranged from 7-35 (M=24.05, SD=5.31). The SDQTD categories were ‘close to average’ (score 0-14): N=106,475 (55.5%), ‘slightly raised’ (score 15-17): N=26,867 (14.0%), ‘high’ (score 18-19): N=15,501 (8.1%), and ‘very high’ (score 20-40): N=43,132 (22.5%).

### Univariable and multivariable multilevel linear regression – SDQTD and SWEMWBS

Table 1 presents the association between SDQTD score and SWEMWBS for 191,975 individuals aged 11-16 in Welsh secondary schools during 2019-2021 using multilevel linear regression. In the unadjusted model (Model 1), the fixed effect intercept was 25.65, indicating that those registering as average on SDQTD experience moderate WB (defined as 21-27 on the SWEWMBS scale). There was evidence of a clear dose-response relationship between SDQTD and SWEMWBS (Table 1). Increasing severity of difficulties (as measured by SDQTD) was associated with worsening wellbeing (as measured by SWEMWBS), and this was consistent after adjustment for demographic (model 2), behavioural (model 3) and social (model 4) covariates.

**Table 1.** Association between Strengths and Difficulties Questionnaire Total Difficulties score (SDQTD) and Short Warwick-Edinburgh Mental Wellbeing Scale (SWEMWBS) for 191,975 individuals aged 11-16 in Welsh Secondary schools from 2019-2021. Intercept, Estimates and their 95% confidence intervals are included for each of the four models.

|  | Unadjusted model<br>(Model 1) |  | Adjusted for demographic variables<br>(Model 2) |  | Adjusted for demographic & behavioural variables<br>(Model 3) |  | Adjusted for demographic, behavioural and social variables<br>(Model 4) |  |
| --- | --- | --- | --- | --- | --- | --- | --- | --- |
|  | *b*<br>(95%CI) | *p* | *b*<br>(95%CI) | *p* | *b*<br>(95%CI) | *p* | *b*<br>(95%CI) | *p* |
| *Intercept* | 25.65<br>(25.59, 25.71) | <2e-16 | 26.33<br>(26.25, 26.41) | <2e-16 | 25.76<br>(25.63, 25.89) | <2e-16 | 26.54<br>(26.41, 26.67) | <2e-16 |
| *SDQ 0-13*<br>*(Close to average)* | Reference group |  | Reference group |  | Reference group |  | Reference group |  |
| *SDQ 14-16*<br>*(Slightly raised)* | -3.26<br>(-3.35, -3.18) | <2e-16 | -2.77<br>(-2.87, -2.66) | <2e-16 | -2.11<br>(-2.19, -2.04) | <2e-16 | -1.61<br>(-1.69, -1.54) | <2e-16 |
| *SDQ 17-19*<br>*(High)* | -4.48<br>(-4.59, -4.37) | <2e-16 | -3.64<br>(-3.76, -3.50) | <2e-16 | -2.99<br>(-3.09, -2.89) | <2e-16 | -2.30<br>(-2.39, -2.21) | <2e-16 |
| *SDQ 20-40*<br>*(Very high)* | -6.67<br>(-6.76, -6.57) | <2e-16 | -5.86<br>(-6.04, -5.68) | <2e-16 | -4.62<br>(-4.72, -4.51) | <2e-16 | -3.42<br>(-3.52, -3.31) | <2e-16 |

Note: Model 1: unadjusted model; Model 2: adjusted for age, sex, and year of assessment; Model 3: further adjusted for tobacco smoking, alcohol consumption, drug use, physical activity, sleep difficulties, and body image; Model 4: further adjusted for family practical support, support from friends, children's living arrangements, having been bullied, school support, care from teachers, and the Family Affluence Scale. SWEMWBS of 21-27 indicates moderate wellbeing. Multiple imputation to address missingness. Higher categories indicate worsening mental health difficulties.

**Table 2.** Within- and between-group variability for the association between Strengths and Difficulties Questionnaire Total Difficulties score (SDQTD) and Short Warwick-Edinburgh Mental Wellbeing Scale (SWEMWBS) for 191,975 individuals across 193 schools (multiple imputed analysis)

|  |  | Variance | SD |
| --- | --- | --- | --- |
| Unadjusted model (Model 1) | School ID | 0.09 | 0.30 |
|  | Residual | 23.61 | 4.86 |
| Adjusted for demographic variables (Model 2) | School ID | 0.09 | 0.30 |
|  | Residual | 22.85 | 4.78 |
| Adjusted for demographic and behavioural variables (Model 3) | School ID | 0.06 | 0.25 |
|  | Residual | 20.69 | 4.55 |
| Adjusted for demographic, behavioural and social variables (Model 4) | School ID | 0.05 | 0.24 |
|  | Residual | 19.19 | 4.38 |

Note: Model 1: unadjusted model; Model 2: adjusted for age, sex, and year of assessment; Model 3: further adjusted for tobacco smoking, alcohol consumption, drug use, physical activity, sleep difficulties, and body image; Model 4: further adjusted for family practical support, support from friends, children's living arrangements, having been bullied, school support, care from teachers, and the Family Affluence Scale. SD: standard deviation

In the fully adjusted model (Model 4), individuals scoring ‘slightly raised’ on the SDQTD were associated with lower SWEMWBS scores (*b* = −1.61, 95%CI = −1.69 to −1.54, p<2e-16) compared to individuals scoring ‘close to average’. This estimate decreased for those scoring ‘high’ (*b* = −2.30, 95%CI = −2.39 to −2.21, p<2e-16), and *‘*very high’ on the SDQTD (*b* = −3.42, 95%CI = −3.52 to −3.31, p<2e-16). In Model 4, the intercept was 26.54, indicating that those registering as ‘close to average’ on SDQTD experience moderate wellbeing (defined as 21-27 on the SWEMWBS scale). Focusing on clinical implications, individuals in with ‘Very high’ severity of difficulties (SDQTD) were associated with a SWEMWBS score of 22, which is near the lower bound of possible mild depression according to the categorical interpretation of SWEMWBS scores(Warwick Medical School, 2021). In terms of the influence of including covariates in the models, the strongest associations were found between the behavioural and social domains in the relationship between SDQTD and SWEMWBS (Supporting Information Table S2).

### Between- and within-group variability

Between-group variation (i.e., variability in school identifier) decreases from Model 1 to Model 4 as part of the variability that initially appeared to be due to differences between schools was explained by the inclusion of covariates. Focusing on the fully adjusted model (Model 4), the random intercept variance between schools is 0.05 (SD=0.24), indicating that the variability in the intercepts across different schools is relatively small, suggesting that the mean outcome (i.e., SWEMWBS scores) does not differ substantially between schools. Conversely, the residual variance representing within-school variability, is 19.19 (SD=4.38). This residual variance implies there is considerable variability in the outcome that is not explained by the differences between schools, but rather by individual differences within schools. The comparison between the random intercept variance and the residual variance highlights that most of the unexplained variability occurs at individual-level rather than at school-level. Results from the between- and within-group variability for models examining the association between each of the SDQ subscales and SWEMWBS (Supporting Information Table S3), and in the complete case model examining the association between the SDQTD and SWEMWBS (Supporting Information Table S4) show a similar pattern to the primary results.

### Secondary analyses

#### Univariable and multivariable multilevel linear regression - SDQ subscales and SWEMWBS

A clear statistically significant dose-response relationship was found for increasing difficulties across each of the SDQ subscales and SWEMWBS (Table 3). The emotional difficulties subscale showed the strongest association with SWEMWBS. In the unadjusted model (Model 1), individuals scoring ‘slightly raised’ in the emotional difficulties were associated with lower SWEMWBS scores (*b* = −2.95, 95%CI = −3.05 to −2.85, p<2e-16) compared to individuals scoring ‘close to average’ SDQ emotional difficulties. This decreased for those with ‘high’ SDQ emotional symptoms (*b* = −3.92, 95%CI = −4.04 to −3.81, p<2e-16) and ‘very high’ SDQ emotional difficulties (*b* = −6.33, 95%CI = −6.49 to −6.17, p<2e-16).

**Table 3.**
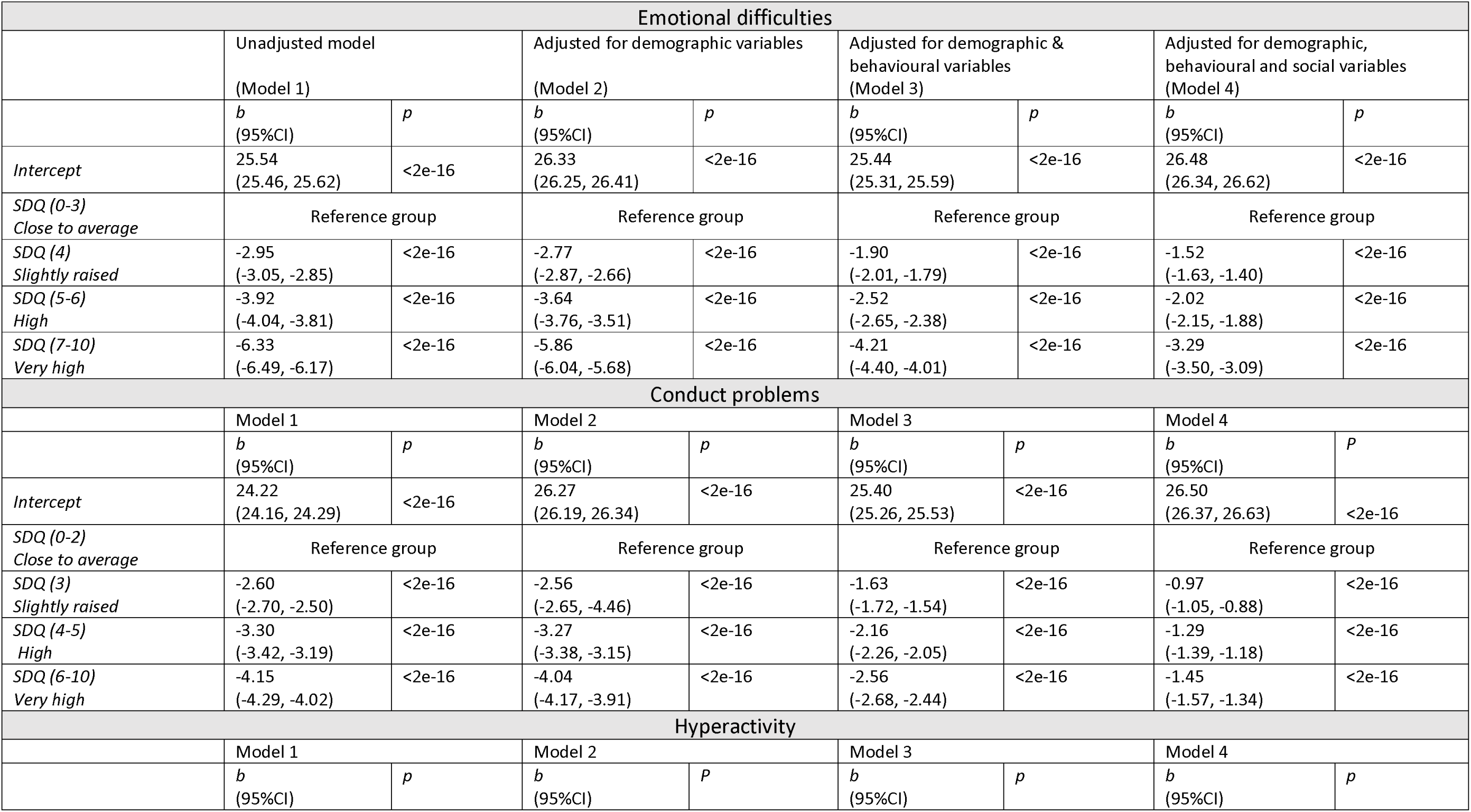
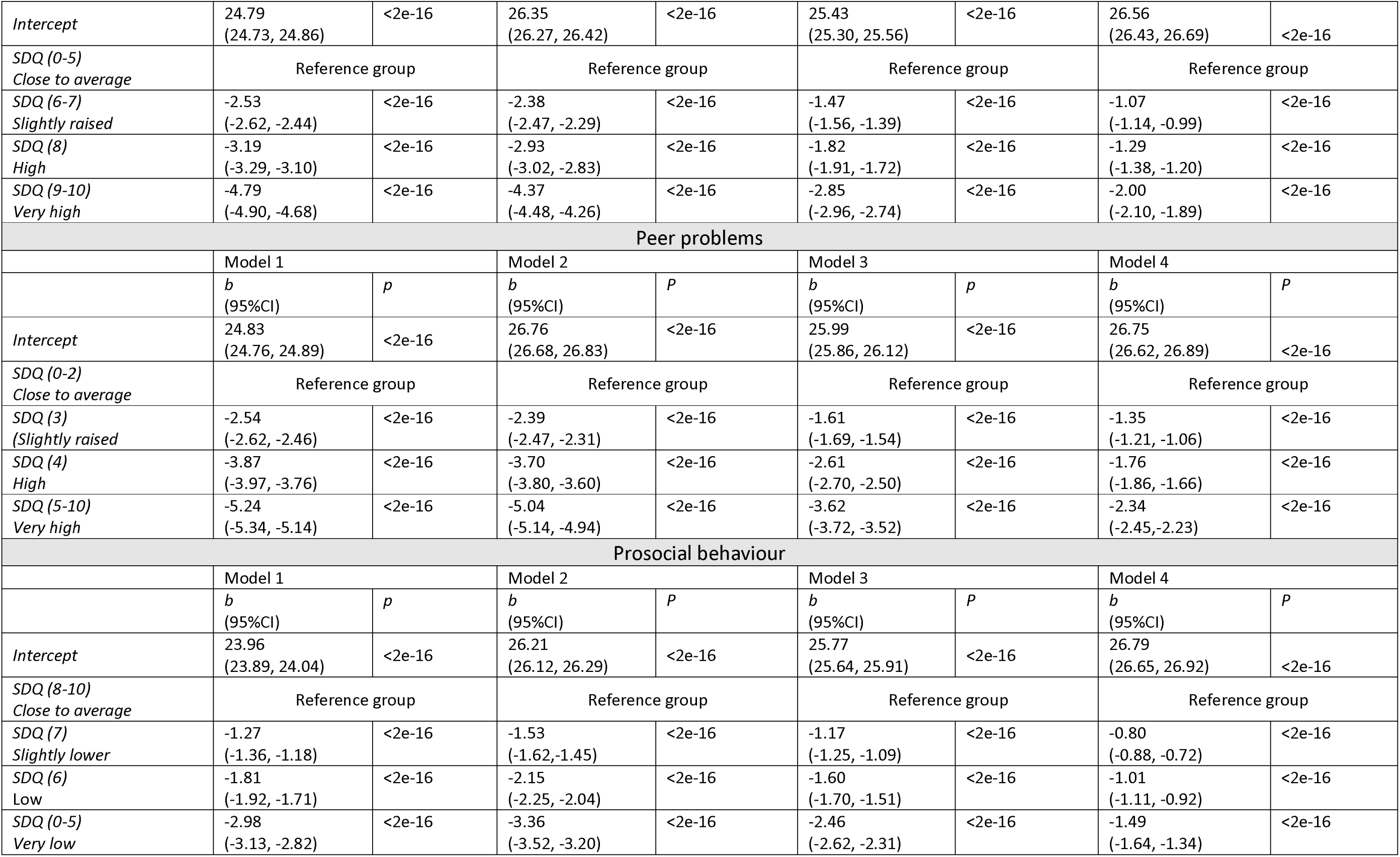
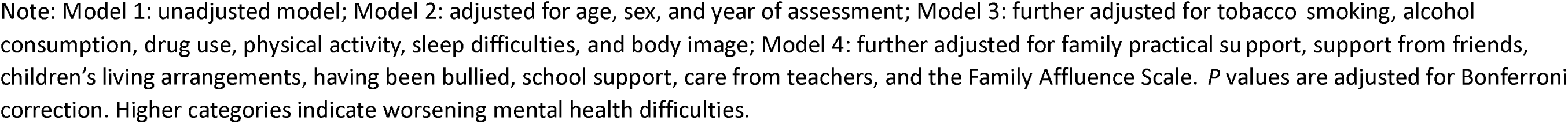
Association between Strengths and Difficulties Questionnaire (SDQ) subscales and Short Warwick-Edinburgh Mental Wellbeing Scale (SWEMWBS) for 191,975 individuals aged 11-16 in Welsh Secondary schools from 2019-2021. Intercept, Estimates and their 95% confidence intervals are included for each of the four models.

| Emotional difficulties |  |  |  |  |  |  |  |  |
| --- | --- | --- | --- | --- | --- | --- | --- | --- |
|  | Unadjusted model<br>(Model 1) |  | Adjusted for demographic variables<br>(Model 2) |  | Adjusted for demographic & behavioural variables<br>(Model 3) |  | Adjusted for demographic, behavioural and social variables<br>(Model 4) |  |
|  | *b*<br>(95%CI) | *p* | *b*<br>(95%CI) | *p* | *b*<br>(95%CI) | *p* | *b*<br>(95%CI) | *p* |
| *Intercept* | 25.54<br>(25.46, 25.62) | <2e-16 | 26.33<br>(26.25, 26.41) | <2e-16 | 25.44<br>(25.31, 25.59) | <2e-16 | 26.48<br>(26.34, 26.62) | <2e-16 |
| *SDQ (0-3)*<br>*Close to average* | Reference group |  | Reference group |  | Reference group |  | Reference group |  |
| *SDQ (4)*<br>*Slightly raised* | -2.95<br>(-3.05, -2.85) | <2e-16 | -2.77<br>(-2.87, -2.66) | <2e-16 | -1.90<br>(-2.01, -1.79) | <2e-16 | -1.52<br>(-1.63, -1.40) | <2e-16 |
| *SDQ (5-6)*<br>*High* | -3.92<br>(-4.04, -3.81) | <2e-16 | -3.64<br>(-3.76, -3.51) | <2e-16 | -2.52<br>(-2.65, -2.38) | <2e-16 | -2.02<br>(-2.15, -1.88) | <2e-16 |
| *SDQ (7-10)*<br>*Very high* | -6.33<br>(-6.49, -6.17) | <2e-16 | -5.86<br>(-6.04, -5.68) | <2e-16 | -4.21<br>(-4.40, -4.01) | <2e-16 | -3.29<br>(-3.50, -3.09) | <2e-16 |
| Conduct problems |  |  |  |  |  |  |  |  |
|  | Model 1 |  | Model 2 |  | Model 3 |  | Model 4 |  |
|  | *b*<br>(95%CI) | *p* | *b*<br>(95%CI) | *p* | *b*<br>(95%CI) | *p* | *b*<br>(95%CI) | *p* |
| *Intercept* | 24.22<br>(24.16, 24.29) | <2e-16 | 26.27<br>(26.19, 26.34) | <2e-16 | 25.40<br>(25.26, 25.53) | <2e-16 | 26.50<br>(26.37, 26.63) | <2e-16 |
| *SDQ (0-2)*<br>*Close to average* | Reference group |  | Reference group |  | Reference group |  | Reference group |  |
| *SDQ (3)*<br>*Slightly raised* | -2.60<br>(-2.70, -2.50) | <2e-16 | -2.56<br>(-2.65, -4.46) | <2e-16 | -1.63<br>(-1.72, -1.54) | <2e-16 | -0.97<br>(-1.05, -0.88) | <2e-16 |
| *SDQ (4-5)*<br>*High* | -3.30<br>(-3.42, -3.19) | <2e-16 | -3.27<br>(-3.38, -3.15) | <2e-16 | -2.16<br>(-2.26, -2.05) | <2e-16 | -1.29<br>(-1.39, -1.18) | <2e-16 |
| *SDQ (6-10)*<br>*Very high* | -4.15<br>(-4.29, -4.02) | <2e-16 | -4.04<br>(-4.17, -3.91) | <2e-16 | -2.56<br>(-2.68, -2.44) | <2e-16 | -1.45<br>(-1.57, -1.34) | <2e-16 |
| Hyperactivity |  |  |  |  |  |  |  |  |
|  | Model 1 |  | Model 2 |  | Model 3 |  | Model 4 |  |
|  | *b*<br>(95%CI) | *p* | *b*<br>(95%CI) | *p* | *b*<br>(95%CI) | *p* | *b*<br>(95%CI) | *p* |
| *Intercept* | 24.79<br>(24.73, 24.86) | <2e-16 | 26.35<br>(26.27, 26.42) | <2e-16 | 25.43<br>(25.30, 25.56) | <2e-16 | 26.56<br>(26.43, 26.69) | <2e-16 |
| *SDQ (0-5)*<br>*Close to average* | Reference group |  | Reference group |  | Reference group |  | Reference group |  |
| *SDQ (6-7)*<br>*Slightly raised* | -2.53<br>(-2.62, -2.44) | <2e-16 | -2.38<br>(-2.47, -2.29) | <2e-16 | -1.47<br>(-1.56, -1.39) | <2e-16 | -1.07<br>(-1.14, -0.99) | <2e-16 |
| *SDQ (8)*<br>*High* | -3.19<br>(-3.29, -3.10) | <2e-16 | -2.93<br>(-3.02, -2.83) | <2e-16 | -1.82<br>(-1.91, -1.72) | <2e-16 | -1.29<br>(-1.38, -1.20) | <2e-16 |
| *SDQ (9-10)*<br>*Very high* | -4.79<br>(-4.90, -4.68) | <2e-16 | -4.37<br>(-4.48, -4.26) | <2e-16 | -2.85<br>(-2.96, -2.74) | <2e-16 | -2.00<br>(-2.10, -1.89) | <2e-16 |
| Peer problems |  |  |  |  |  |  |  |  |
|  | Model 1 |  | Model 2 |  | Model 3 |  | Model 4 |  |
|  | *b*<br>(95%CI) | *p* | *b*<br>(95%CI) | *p* | *b*<br>(95%CI) | *p* | *b*<br>(95%CI) | *p* |
| *Intercept* | 24.83<br>(24.76, 24.89) | <2e-16 | 26.76<br>(26.68, 26.83) | <2e-16 | 25.99<br>(25.86, 26.12) | <2e-16 | 26.75<br>(26.62, 26.89) | <2e-16 |
| *SDQ (0-2)*<br>*Close to average* | Reference group |  | Reference group |  | Reference group |  | Reference group |  |
| *SDQ (3)*<br>*(Slightly raised)* | -2.54<br>(-2.62, -2.46) | <2e-16 | -2.39<br>(-2.47, -2.31) | <2e-16 | -1.61<br>(-1.69, -1.54) | <2e-16 | -1.35<br>(-1.21, -1.06) | <2e-16 |
| *SDQ (4)*<br>*High* | -3.87<br>(-3.97, -3.76) | <2e-16 | -3.70<br>(-3.80, -3.60) | <2e-16 | -2.61<br>(-2.70, -2.50) | <2e-16 | -1.76<br>(-1.86, -1.66) | <2e-16 |
| *SDQ (5-10)*<br>*Very high* | -5.24<br>(-5.34, -5.14) | <2e-16 | -5.04<br>(-5.14, -4.94) | <2e-16 | -3.62<br>(-3.72, -3.52) | <2e-16 | -2.34<br>(-2.45, -2.23) | <2e-16 |
| Prosocial behaviour |  |  |  |  |  |  |  |  |
|  | Model 1 |  | Model 2 |  | Model 3 |  | Model 4 |  |
|  | *b*<br>(95%CI) | *p* | *b*<br>(95%CI) | *p* | *b*<br>(95%CI) | *p* | *b*<br>(95%CI) | *p* |
| *Intercept* | 23.96<br>(23.89, 24.04) | <2e-16 | 26.21<br>(26.12, 26.29) | <2e-16 | 25.77<br>(25.64, 25.91) | <2e-16 | 26.79<br>(26.65, 26.92) | <2e-16 |
| *SDQ (8-10)*<br>*Close to average* | Reference group |  | Reference group |  | Reference group |  | Reference group |  |
| *SDQ (7)*<br>*Slightly lower* | -1.27<br>(-1.36, -1.18) | <2e-16 | -1.53<br>(-1.62, -1.45) | <2e-16 | -1.17<br>(-1.25, -1.09) | <2e-16 | -0.80<br>(-0.88, -0.72) | <2e-16 |
| *SDQ (6)*<br>*Low* | -1.81<br>(-1.92, -1.71) | <2e-16 | -2.15<br>(-2.25, -2.04) | <2e-16 | -1.60<br>(-1.70, -1.51) | <2e-16 | -1.01<br>(-1.11, -0.92) | <2e-16 |
| *SDQ (0-5)*<br>*Very low* | -2.98<br>(-3.13, -2.82) | <2e-16 | -3.36<br>(-3.52, -3.20) | <2e-16 | -2.46<br>(-2.62, -2.31) | <2e-16 | -1.49<br>(-1.64, -1.34) | <2e-16 |

Note: Model 1: unadjusted model; Model 2: adjusted for age, sex, and year of assessment; Model 3: further adjusted for tobacco smoking, alcohol consumption, drug use, physical activity, sleep difficulties, and body image; Model 4: further adjusted for family practical support, support from friends, children's living arrangements, having been bullied, school support, care from teachers, and the Family Affluence Scale. *P* values are adjusted for Bonferroni correction. Higher categories indicate worsening mental health difficulties.

#### Sensitivity analysis

Sensitivity analyses were included for both the primary analyses examining the relationship between the SDQTD and SWEMWBS, and for the secondary analyses examining the relationship between the SDQ subscales and SWEMWBS: i) complete case sample (Supporting Information Tables S5 and S6), ii) sample that consisted of individuals who had incomplete information on SWEMWBS, SDQTD, and covariates (Supporting Information Tables S7and S8), iii) 2019/20 year of assessment (Supporting Information Tables S9 and S10), and iv) 2021/2021 year of assessment (Supporting Information Tables S11 and S12). Overall, the findings remained consistent to the main analyses across all sensitivity samples.

## DISCUSSION

### Summary of results

In this study, we found strong evidence of a dose-response relationship between MH difficulties and WB in a nationally representative population of over 190,000 pupils aged 11-16 years in Wales. Increasing MH difficulties were associated with worsening WB and remained after adjusting for demographic, behavioural, and social variables. The same dose-response relationships were evident when examining the association between the specific MH subscales (i.e., emotional problems, conduct problems, hyperactivity, and peer problems) and WB. This pattern of results was consistent across multiple sensitivity analysis including complete cases, multiple imputed samples, and by analysing different cohort years separately. Variation between schools was small, but there were high levels of within-school variance, highlighting the important differences between individuals in terms of their MH and WB.

### Comparison with previous literature

Previous studies examining the relationship between MH and WB acknowledge a complex interrelated association, while also showing an inconsistency in defining and measuring the two constructs. Many studies consistently show that MH and WB are interconnected but distinct constructs (Deighton et al., 2019; Keyes, 2005; Patalay & Fitzsimons, 2016a, 2018; Suldo & Shaffer, 2008), while others demonstrate evidence the MH and WB are part of a single overarching construct (Gerhardt et al., 2021; Trudel-Fitzgerald et al., 2019). Our findings identified a dose-response relationship between increasing MH difficulties and worsening WB, which was consistent after adjusting for other covariates, and within sub-domains of SDQ. Our study supports the argument that MH and WB are discrete constructs and whilst there is a clear quantifiable relationship between them, they are distinct. This suggests that addressing both constructs simultaneously may be more effective than prioritising one over the other. Our study also demonstrates the strong association, specifically within the emotional MH subscale and WB. In this regard, our findings strengthen the argument that improving MH is beneficial to WB and vice versa (Viner et al., 2022).

Our findings indicate that schools were not the primary source of variation in MH or WB between individuals. After adjusting for individual and family differences across socioeconomic background, various behavioural factors, and indicators reflecting support systems, there was little remaining evidence of specific school-level effects. This was not expected given that school environments have been reported to contribute to MH and WB experiences and play a significant role in MH and WB (Ravens-Sieberer, Erhart, Gosch, Wille, & Group, 2008). Adjusting for covariates at the individual level may have accounted for some school effects. For instance, if a trusted adult was a teacher rather than a parent, or if there were experiences of bullying, these factors could have been captured in the individual-level adjustments. Consequently, this might have resulted in no observable school-level effect.

Schools are often the target of settings-based interventions to address the underlying causes and to support MH and WB, but our study emphasises that individual-level factors play a substantial role on MH and WB outcomes. These findings potentially highlight the importance of personalised interventions and support systems tailored to the unique needs of each student. While they could also highlight the importance of a proportionate universalism perspective (Marmot, 2010). Whereby interventions could be designed to benefit everyone, but with varying levels of intensity based on need, ensures that resources are allocated in a way that maximises impact and reduces inequalities.

### Strengths and Limitations

A key strength of this study was to use a nationally representative sample of Welsh schoolchildren aged 11-16 years, unlike the sample used in (Lereya et al., 2022) that focused on disadvantaged areas. This approach helped to minimise the possibility of Type I and Type II error. Data in this study was treated as whole-population sampling as randomisation of the 95% of Welsh schools participated in the SHRN survey was performed at a national-level rather than school-level. This allows for a more generalisable understanding of the MH and WB of adolescents within this age group. Furthermore, the use of self-report MH and WB measures in this study unlike the approach used in (Patalay & Fitzsimons, 2016b), which has been shown to be as they capture internal experiences (Ravens-Sieberer et al., 2008). Using a nationally representative sample that includes comprehensive data on demographic, behavioural, and social confounders at individual, family and household levels, addresses common limitations in previous research (e.g., using different raters for young people MH and WB, failing to control for all potential confounding domains, and not utilising a nationally representative sample).

The present study should be considered in light of some limitations. Although, there is potential overlap between the MH and mental WB tools used, these tools are well-validated and widely used in research, providing a robust framework for assessing MH and WB. The SDQ and the SWEMWBS have been extensively tested for reliability and validity, ensuring that they accurately measure the constructs of interest. Furthermore, the overlap between tools can be seen as a reflection of the interconnected nature of MH and WB. Emotional difficulties, for example, are inherently linked to overall wellbeing, and capturing this overlap can provide a more holistic understanding of an individual’s mental state (Keyes, 2005).

Second, although participants are expected to answer all questions, some may skip or omit certain items, leading to missing data. We attempted to estimate the maximum effect of attrition using sensitivity analyses of complete cases and multiple imputations samples. The results from the sensitivity analysis suggest that the pattern of missing data did not lead to biased effect estimates. Third, although this study is nationally representative, it is important to acknowledge that, like all studies, it cannot control for every possible confounder. There may still be unmeasured variables that influence the relationship between MH and WB in young people. Studies have shown a strong genetic overlap between traits like neuroticism, anxiety, and depression, which are key contributors to MH issues, and these same genetic factors can influence overall wellbeing (Nivard et al., 2015). Finally, the categorisation of SDQ in this study rather than the use of raw SDQ scores. This prevented the examination of internalising and externalising scales and prevented higher resolution examination of the location and spread of scores by group in Table 1.

### Conclusion

These results demonstrated a clear and consistent association between increasing MH difficulties and worsening MW, and that these are two discrete constructs with interdependencies. This study underscores the need for an integrated framework that considers the interplay between MH and WB, ensuring that prevention of poor MH and WB, and protection of good MH and WB is holistic and addresses the full spectrum of adolescent needs. By bridging the gap between these constructs, researchers and practitioners can develop more robust and effective programs that promote overall adolescent development and resilience.

## Data Availability

The data that support the findings of this study are available from the School Health Research Network (SHRN). Interested researchers must submit a research proposal to SHRN. For more information on how to apply, please see https://www.shrn.org.uk/

## Acknowledgements

The authors are grateful for the pupils who participated in the survey.

## Notes

### Competing Interest Statement

The authors have declared no competing interest.

### Funding Statement

This study was funded by Public Health Wales NHS Trust; Wolfson Foundation; Medical Research Council.

### Author Declarations

Ethical approval for Student Health and Well-being surveys (2017-23) was obtained from Cardiff University's School of Social Sciences Research Ethics Committee. Details regarding the ethical considerations and approvals for this study are available here (https://doi.org/10.1093/ije/dyae161).

