## Supporting Information for "Investigating the relationship between adolescent mental health and wellbeing in a nationally representative survey of 11-16 year olds in Wales"

**Supplementary Material**

**Table S1**. Association between covariates and non-availability of SWEMWBS data

**Table S2.** Descriptive information of each covariate mental wellbeing (multiple imputed analysis N=191,975)

**Table S3.** Within- and between-group variability for the association between each of the SDQ subscales and SWEMWBS

**Table S4.** Within- and between-group variability for the association between SDQTD score and SWEMWBS for 114,439 individuals across 193 schools (complete case analysis)

**Table S5.** Association between SDQTD score and SWEMWBS for 114,439 individuals aged 11-16 in Welsh Secondary schools from 2019-2021. Intercept, Estimates and their 95% confidence intervals are included for each of the 4 models.

**Table S6.** Association between SDQ subscales and SWEMWBS for 114,439 individuals aged 11-16 in Welsh Secondary schools from 2019-2021. Intercept, Estimates and their 95% confidence intervals are included for each of the 4 models.

**Table S7.** Association between SDQTD score and SWEMWBS for 212,975 individuals aged 11-16 in Welsh Secondary schools from 2019-2021. Intercept, Estimates and their 95% confidence intervals are included for each of the 4 models.

**Table S8.** Association between SDQ subscales and SWEMWBS for 212,975 individuals aged 11-16 in Welsh Secondary schools from 2019-2021. Intercept, Estimates and their 95% confidence intervals are included for each of the 4 models.

**Table S9.** Association between SDQTD score and SWEMWBS for 105,934 individuals aged 11-16 in Welsh Secondary schools from 2019-2021. Intercept, Estimates and their 95% confidence intervals are included for each of the 4 models (2019 survey).

**Table S10.** Association between SDQ subscales and SWEMWBS for 105,934 individuals aged 11-16 in Welsh Secondary schools from 2019-2021. Intercept, Estimates and their 95% confidence intervals are included for each of the 4 models (2019 survey).

**Table S11.** Association between SDQTD score and SWEMWBS for 109,031 individuals aged 11-16 in Welsh Secondary schools from 2019-2021. Intercept, Estimates and their 95% confidence intervals are included for each of the 4 models (2021 survey).

**Table S12.** Association between SDQ subscales and SWEMWBS for 109,031 individuals aged 11-16 in Welsh Secondary schools from 2019-2021. Intercept, Estimates and their 95% confidence intervals are included for each of the 4 models (2019 survey).

***Covariates***

*Age*

Age was calculated using given and derived variables. Children supplied their own birth year and month as survey answers. The survey month and year were converted into months elapsed since the year 0, as were the birth month and year. The difference between these month counts was divided by 12 to provide an estimated age in years. These ages were grouped into categories of two years each: 11-12 years, 13-14 years and 15-16 years. Children with calculated ages < 11 years (n=141) or > 16 years (n=173) were excluded from the dataset.

*Sex*

In 2019 and 2021, when asked about their sex, children were able to answer ‘Male’, ‘Female’ or ‘Neither word describes me’. The last category was interpreted here as ‘Non-binary’. In 2017, the possible options for this question were simply “Boy” or “Girl”.

*Year of assessment*

Year of completion of survey (i.e., 2019 or 2021) was included.

*Smoking*

Children were asked how often they smoked cigarettes on a 4-point scale. These responses were simplified into a binary variable: “Every day” or “At least once a week” was classed as “Yes”, while “Less than once a week” or “I do not smoke” was classed as “No”.

*Alcohol consumption*

Children were asked how often they drank beer, wine, spirits, cider, alcopops or other alcohol. Responses to these six questions were aggregated to create a single variable for frequency of alcohol consumption. The categories for this variable were simplified from a 5-point scale to “Never”, “Less than weekly” and “Weekly or more”.

*Drug use*

Children were asked how recently they had used laughing gas, mephedrone and legal highs. They were also asked about cannabis use in the last 30 days and in their lifetime. Responses to these five questions were aggregated into a single variable depicting recency of drug use: “Never”, “> 30 days ago” and “<= 30 days ago”.

*Physical activity*

Children were asked to recount the number of days out of the last seven on which they had been physically active for at least 60 minutes. Their responses were simplified from an 8-point scale ranging from 0 days to 7 days into “Inactive” (0 days), “Less active” (1-3 days) and “More active” (4-7 days).

*Sleep difficulty*

Children were asked how often they had difficulty sleeping in the last six months. Their responses were simplified from a 5-point frequency scale into a binary variable in which individuals reporting sleeping difficulty every week or more frequently were classed as ‘Yes’, while those who experienced sleeping difficulty monthly or less frequently were classed as ‘No’.

*Body image*

Children were asked how they considered their own body size. Their responses were simplified from a 5-point scale ranging from ‘Much too thin’ to ‘Much too fat’ into a three-category variable comprising ‘Too thin’, ‘About right’ and ‘Too fat’.

*Lives with*

Children answered a series of binary questions concerning with whom or where they lived: with their mother, father, stepmother, stepfather, grandparents, aunt or uncle, sibling, or foster parents, in a residential home, independently or elsewhere. These were combined into one variable which described whether they lived with both parents, one parent, with a non-parent relative, in care (foster or residential) or elsewhere (independently or elsewhere). The assumption of a heteronormative familial structure was unavoidable due to limitations in the question structure: children living with two mothers, or two fathers will incorrectly be assigned to the “One parent” category.

*Family support and support from friends*

Children were asked whether their family really tried to help them. Their responses were simplified from 7-point scales (‘Very strongly disagree’ to ‘Very strongly agree’) into ‘No’ (1-3), ‘Ambivalent’ (4) and ‘Yes’ (5-7). Their perceived ability to count on their friends was treated similarly.

*Bullying others*

Children were asked how often they had enacted or experienced bullying in the last two months. Responses to each of these questions were simplified into “Never”, “Less than weekly” and “At least weekly”.

*School support and teachers caring*

Children were asked whether they felt their teachers cared about them whether there was support for pupils who were unhappy or not coping. Their responses were simplified from 5-point scales (‘Strongly disagree’ to ‘Strongly agree’) into ‘No’ (1-2), ‘Ambivalent’ (3) and ‘Yes’ (4-5).

*Family Affluence Scale (FAS)*

The Family Affluence Scale was used as a measure of socioeconomic status (Currie et al., 1997). The scale comprises six questions about bedroom-sharing, domestic bathroom count, holidaymaking and computer, dishwasher and car ownership. Given significant changes in the pattern of holidaymaking in 2021 due to the COVID-19 pandemic, this question was dropped from the scale and a 5-item scale was used in all years. Scores were presented as tertiles.

*Exposure*

*Strengths and Difficulties Questionnaire (SDQ)*

SDQ’s validation is against the Rutter parents’ and teachers’ scale (Goodman, 1997), a screening tool for behavioral problems, which was designed to assess changes in “disturbed behaviour” over time (Elander & Rutter, 1996).SDQ is particularly useful in screening for neurodivergence such as attention deficit hyperactivity disorder (sensitivity 0.91, specificity 0.90) and autism spectrum disorder (sensitivity 0.79, specificity 0.93) (Russell et al., 2013). Concurrent criterion validity was assessed insofar as it was found to be able to distinguish between paediatric dental and psychiatric populations (Goodman, 1997).

Each is assessed by asking the child to respond, “Not true”, “Somewhat true” or “Certainly true” to five statements in each scale. For the four ‘problems’ scales, statements about undesirable traits, such as “I worry a lot” (emotional problems) are scored as “Certainly true” = 2, “Somewhat true” = 1 and “Not true” = 0. Statements about desirable traits, such as “Other people my age generally like me” (peer problems), are scored in reverse, so “Certainly true” = 0, “Somewhat true” = 1 and “Not true” = 2. The final ‘prosocial behaviour’ scale is treated differently. All statements refer to desirable traits, such as “I am helpful if someone is hurt” and are scored “Certainly true” = 2, “Somewhat true” = 1 and “Not true” = 0. In this study, raw scores were not available.

*Outcome*

*Short Warwick-Edinburgh Mental Wellbeing Scale (SWEMWBS)*

A secondary analysis of 13 studies which used the full WEMWBS found the tool to be sensitive to changes in WB (Maheswaran et al., 2012). While the majority of studies validating the tool have focused on WEMWBS, an appraisal specifically of SWEMWBS found that scores were moderately correlated with indicators of life satisfaction and somatisation (Melendez-Torres et al., 2019). SWEMWBS has also been found to be more precise than the original due to its scaling properties (Stewart-Brown et al., 2009). Despite its intended purpose being to monitor the general population, SWEMWBS has primarily been validated in clinical populations (Shah et al., 2018, 2021).

Table S1. Association between covariates and non-availability of SWEMWBS data

|  |  | OR | 95% CI  low | 95% CI high | p |
| --- | --- | --- | --- | --- | --- |
| Age | 11-12 years | reference | | | |
|  | 13-14 years | 0.55 | 0.50 | 0.60 | <2e-16 |
|  | 15-16 years | 0.32 | 0.28 | 0.36 | <2e-16 |
| Sex | Male | reference | | | |
|  | Female | 1.14 | 1.06 | 1.23 | 0.000618 |
|  | Non-binary | 1.03 | 0.76 | 1.35 | 0.863836 |
| Smoking | No | reference | | | |
|  | Yes | 1.26 | 0.97 | 1.63 | 0.080948 |
| Alcohol consumption | Never | reference | | | |
|  | Up to 1 drink | 0.81 | 0.73 | 0.90 | 0.00011 |
|  | 2-3 drinks | 0.81 | 0.69 | 0.94 | 0.005758 |
|  | 4+ drinks | 0.79 | 0.66 | 0.95 | 0.01459 |
| Drug use | Never | reference | | | |
|  | > 30 days ago | 1.41 | 0.79 | 2.36 | 0.213986 |
|  | <= 30 days ago | 0.99 | 0.53 | 1.73 | 0.972935 |
| Physical activity | Inactive | reference | | | |
|  | Less active | 0.91 | 0.76 | 1.10 | 0.338369 |
|  | More active | 0.94 | 0.78 | 1.13 | 0.500483 |
| Sleep difficulty | No | reference | | | |
|  | Yes | 0.97 | 0.89 | 1.05 | 0.431407 |
| Body image | Too thin | reference | | | |
|  | About right | 1.10 | 0.98 | 1.23 | 0.09802 |
|  | Too fat | 0.99 | 0.90 | 1.08 | 0.760947 |
| Lives with | Two parents | reference | | | |
|  | One parent | 1.00 | 0.92 | 1.09 | 0.939858 |
|  | Relative | 1.44 | 1.10 | 1.86 | 0.006185 |
|  | Care | 1.65 | 1.07 | 2.43 | 0.015789 |
|  | Elsewhere | 1.69 | 0.84 | 3.09 | 0.109772 |
| Family practical | Yes | reference | | | |
|  | Ambivalent | 0.84 | 0.71 | 0.99 | 0.042264 |
|  | No | 0.96 | 0.86 | 1.07 | 0.498307 |
| Friends support | Yes | reference | | | |
|  | Ambivalent | 1.03 | 0.91 | 1.17 | 0.626251 |
|  | No | 1.10 | 1.00 | 1.20 | 0.053875 |
| Been bullied | Never | reference | | | |
|  | Less than weekly | 1.05 | 0.96 | 1.15 | 0.246053 |
|  | At least weekly | 1.19 | 1.03 | 1.36 | 0.015016 |
| School support | Yes | reference | | | |
|  | Ambivalent | 1.03 | 0.94 | 1.14 | 0.519871 |
|  | No | 1.03 | 0.90 | 1.17 | 0.677657 |
| Teachers care | Yes | reference | | | |
|  | Ambivalent | 1.10 | 1.00 | 1.21 | 0.042998 |
|  | No | 1.21 | 1.08 | 1.37 | 0.00123 |
| Family Affluence Scale | Lowest | reference | | | |
|  | Middle | 0.87 | 0.80 | 0.96 | 0.003284 |
|  | Highest | 0.72 | 0.65 | 0.79 | 1.59e-11 |

*Dealing with missing data*

Multiple logistic regression was used to examine how each of the included variables was related to the likelihood of the outcome being missing (i.e., non-completion of the SWEMWBS). MI was based on 191,975 individuals who provided complete information on SWEMWBS at either of the 2019/20 or 2021/22 surveys and who had incomplete information on the exposure and covariates. As the model contained multiple variables from demographic, behavioural, and social domains, it was deemed unnecessary to include other variables as auxiliary variables in the imputation model. Missing data were handled using the ‘mice’ package in R which utilises chained equations. Continuous variables were imputed using predictive mean matching; numeric variables were imputed using binary logistic regression or polytomous logistic regression. Clustering by school was included in the imputation process. Due to the difficulty in including low cell counts in the MI process, two schools with fewer than 100 students were excluded from the analysis. Imputed datasets were pooled using Rubin’s rules (Rubin, 1987).

Table S2. Descriptive information of each covariate mental wellbeing (multiple imputed analysis N=191,975)

| **Variable** | **Category** | **n** | **%** | **SWEMWBS**  **(Mean)** | **SWEMWBS**  **(SD)** |
| --- | --- | --- | --- | --- | --- |
| **Year** | *2019* | 82989 | 43.2 | 23.7 | 5.59 |
|  | *2021* | 108986 | 56.8 | 23.0 | 5.61 |
| **Age** | *11-12 years* | 69278 | 36.1 | 23.9 | 5.59 |
|  | *13-14 years* | 78503 | 40.9 | 23.2 | 5.61 |
|  | *15-16 years* | 44194 | 23.0 | 22.7 | 5.56 |
| **Sex** | *Male* | 94810 | 49.4 | 24.5 | 5.43 |
|  | *Female* | 92987 | 48.4 | 22.4 | 5.45 |
|  | *Non-binary* | 4178 | 2.2 | 18.1 | 6.01 |
| **Local Authority** | *Swansea* | 14480 | 7.5 | 23.6 | 5.56 |
|  | *Blaenau Gwent* | 3207 | 1.7 | 23.1 | 5.52 |
|  | *Vale of Glamorgan* | 9642 | 5.0 | 24.1 | 5.64 |
|  | *Cardiff* | 18632 | 9.7 | 23.6 | 5.61 |
|  | *Caerphilly* | 12217 | 6.4 | 23.3 | 5.58 |
|  | *Newport* | 10870 | 5.7 | 23.5 | 5.65 |
|  | *Neath Port Talbot* | 10875 | 5.7 | 23.2 | 5.65 |
|  | *Ceredigion* | 4846 | 2.5 | 23.3 | 5.57 |
|  | *Conwy* | 7790 | 4.1 | 23.0 | 5.56 |
|  | *Gwynedd* | 7630 | 4.0 | 23.6 | 5.58 |
|  | *Merthyr Tydfil* | 3236 | 1.7 | 23.0 | 5.73 |
|  | *Bridgend* | 9118 | 4.7 | 23.3 | 5.72 |
|  | *Powys* | 7777 | 4.0 | 23.3 | 5.51 |
|  | *Rhondda Cynon Taf* | 12969 | 6.8 | 23.3 | 5.69 |
|  | *Denbighshire* | 7429 | 3.9 | 23.3 | 5.55 |
|  | *Monmouthshire* | 5580 | 2.9 | 23.4 | 5.53 |
|  | *Carmarthenshire* | 10125 | 5.3 | 23.3 | 5.30 |
|  | *Pembrokeshire* | 7327 | 3.8 | 23.1 | 5.55 |
|  | *Flintshire* | 10870 | 5.7 | 23.0 | 5.60 |
|  | *Torfaen* | 6301 | 3.3 | 23.0 | 5.63 |
|  | *Wrexham* | 7330 | 3.8 | 22.9 | 5.63 |
|  | *Isle of Anglesey* | 3741 | 1.9 | 23.3 | 5.88 |
| **SDQ total difficulties** | *Close to average* | 106475 | 56.6 | 25.6 | 4.74 |
|  | *Slightly raised (15-17)* | 26867 | 13.8 | 22.4 | 4.60 |
|  | *High (18-19)* | 15501 | 7.7 | 21.2 | 4.73 |
|  | *Very high (20-40)* | 43132 | 21.9 | 19.0 | 5.38 |
| **SDQ emotional difficulties** | *Close to average* | 106045 | 55.6 | 25.6 | 4.93 |
|  | *Slightly raised (5)* | 21640 | 11.3 | 22.6 | 4.73 |
|  | *High (5)* | 18615 | 9.7 | 21.6 | 4.75 |
|  | *Very high (7-10)* | 45674 | 23.4 | 19.2 | 5.11 |
| **SDQ conduct problems** | *Close to average* | 139433 | 73.2 | 24.2 | 5.31 |
|  | *Slightly raised (4)* | 20669 | 10.6 | 21.7 | 5.39 |
|  | *High (5)* | 15162 | 7.5 | 20.9 | 5.50 |
|  | *Very high (6-10)* | 16711 | 8.6 | 19.9 | 6.09 |
| **SDQ hyperactivity** | *Close to average* | 115807 | 60.6 | 24.8 | 5.15 |
|  | *Slightly raised (6)* | 22793 | 12.0 | 22.2 | 5.34 |
|  | *High (7)* | 18588 | 9.6 | 21.6 | 5.22 |
|  | *Very high (8-10)* | 34788 | 17.9 | 19.9 | 5.55 |
| **SDQ peer problems** | *Close to average* | 116486 | 61.3 | 24.8 | 5.06 |
|  | *Slightly raised (3)* | 29261 | 15.2 | 22.4 | 5.17 |
|  | *High (4)* | 20656 | 10.8 | 21.1 | 5.72 |
|  | *Very high (5-10)* | 25573 | 12.8 | 19.4 | 5.79 |
| **SDQ prosocial behaviour** | *Close to average (7-10)* | 128169 | 66.8 | 24.0 | 5.46 |
|  | *Slightly raised (6)* | 25028 | 13.1 | 22.8 | 5.33 |
|  | *High (5)* | 18509 | 9.6 | 22.2 | 5.45 |
|  | *Very high (0-4)* | 20270 | 10.5 | 21.1 | 6.22 |
| **Smoking** | *No* | 185447 | 96.5 | 23.5 | 5.51 |
|  | *Yes* | 6528 | 3.5 | 19.1 | 6.77 |
| **Alcohol consumption** | *Never* | 112526 | 58.6 | 24.1 | 5.50 |
|  | *Up to 1 drink* | 37207 | 19.4 | 22.9 | 5.27 |
|  | *2-3 drinks* | 21466 | 11.1 | 22.2 | 542 |
|  | *4+ drinks* | 20776 | 10.9 | 21.1 | 6.01 |
| **Drug use** | *Never* | 189964 | 98.9 | 23.4 | 5.55 |
|  | *> 30 days ago* | 947 | 0.5 | 18.6 | 6.80 |
|  | *<= 30 days ago* | 1064 | 0.6 | 17.3 | 8.79 |
| **Physical activity** | *Inactive* | 9845 | 5.2 | 19.5 | 6.20 |
|  | *Less active* | 65000 | 33.9 | 22.3 | 5.34 |
|  | *More active* | 117131 | 60.9 | 24.3 | 5.46 |
| **Sleep difficulty** | *No* | 100140 | 52.2 | 25.4 | 5.03 |
|  | *Yes* | 91835 | 47.8 | 21.1 | 5.39 |
| **Body image** | *About right* | 96872 | 50.4 | 25.0 | 5.15 |
|  | *Too thin* | 26304 | 13.7 | 22.6 | 5.53 |
|  | *Too fat* | 68799 | 35.8 | 21.3 | 5.53 |
| **Lives with** | *Both parents* | 130614 | 68.5 | 23.8 | 5.48 |
|  | *One parent* | 55966 | 28.1 | 22.3 | 5.63 |
|  | *Relative* | 3437 | 2.0 | 21.5 | 6.20 |
|  | *Care* | 1335 | 0.8 | 21.7 | 6.49 |
|  | *Elsewhere* | 623 | 0.6 | 21.2 | 7.16 |
| **Family practical** | *Yes* | 144837 | 75.6 | 24.2 | 5.20 |
|  | *Ambivalent* | 14090 | 7.2 | 20.1 | 4.89 |
|  | *No* | 33048 | 17.3 | 20.8 | 6.30 |
| **Friends support** | *Yes* | 123209 | 64.4 | 24.2 | 5.36 |
|  | *Ambivalent* | 21585 | 11.1 | 21.7 | 5.05 |
|  | *No* | 47182 | 24.5 | 21.5 | 6.11 |
| **Been bullied** | *No* | 128380 | 67.1 | 24.2 | 5.36 |
|  | *Less than weekly* | 46675 | 24.3 | 22.1 | 5.41 |
|  | *At least weekly* | 16920 | 8.6 | 19.8 | 6.02 |
| **School support** | *Yes* | 122413 | 64.0 | 24.5 | 5.30 |
|  | *Ambivalent* | 44168 | 23.0 | 22.1 | 5.21 |
|  | *No* | 25395 | 13.1 | 19.9 | 5.97 |
| **Teachers care** | *Yes* | 104512 | 54.6 | 24.8 | 5.23 |
|  | *Ambivalent* | 53807 | 28.0 | 22.3 | 5.07 |
|  | *No* | 33656 | 17.4 | 20.4 | 6.05 |
| **Family Affluence Scale** | *Lowest* | 53362 | 27.8 | 22.4 | 5.74 |
|  | *Middle* | 70514 | 36.7 | 23.4 | 5.55 |
|  | *Highest* | 68099 | 35.5 | 24.1 | 5.47 |

*Within- and between-group variability for the association between each of the SDQ subscales and SWEMWBS*

There is a decrease in variance explained due to the inclusion of covariates in models examining the association between all of the SDQ subscales and SWEMWBS resulting in model fit improvements (Table S3). Between-group variation (i.e., variability in school ID) decreases from Model 1 to Model 4 in all SDQ subscales as part of the variability that was initially considered to be due to differences between schools in now explained by the inclusion of covariates. Focusing on models fully adjusted for covariates in all SDQ subscales (Model 4), the random intercept variance between schools is 0.06 (SD=0.23 – 0.25), indicating that the variability in the intercepts across different schools is relatively small, suggesting that the mean outcome (i.e., SWEMWBS scores) does not differ substantially between schools when examining individual SDQ subscales as the exposure. Conversely, the residual variance representing within-school variability, is 19.20 – 20.27 (SD=4.38 – 4.50). This residual variance implies that there is considerable variability in the outcome that is not explained by the differences between schools, but rather by individual differences within schools. The comparison between the random intercept variance and the residual variance highlights that the majority of the unexplained variability occurs at individual level rather than at school level.

Table S3. Within- and between-group variability for the association between each of the SDQ subscales and SWEMWBS

|  |  | Emotional difficulties | | Conduct problems | | Hyperactivity | | Peer problems | | Prosocial behaviour | |
| --- | --- | --- | --- | --- | --- | --- | --- | --- | --- | --- | --- |
|  |  | Variance | SD | Variance | Variance | SD | SD | Variance | SD | Variance | SD |
| Model 1 | School ID | 0.12 | 0.35 | 0.18 | 0.43 | 0.14 | 0.37 | 0.15 | 0.39 | 0.20 | 0.45 |
|  | Residual | 24.22 | 4.92 | 28.99 | 5.38 | 27.57 | 5.25 | 27.29 | 5.22 | 30.21 | 5.49 |
| Model 2 | School ID | 0.13 | 0.35 | 0.16 | 0.39 | 0.14 | 0.37 | 0.13 | 0.37 | 0.17 | 0.41 |
|  | Residual | 23.83 | 4.88 | 27.01 | 5.20 | 26.17 | 5.11 | 25.58 | 5.05 | 27.88 | 5.28 |
| Model 3 | School ID | 0.08 | 0.28 | 0.08 | 0.29 | 0.08 | 0.28 | 0.07 | 0.27 | 0.07 | 0.27 |
|  | Residual | 21.20 | 4.60 | 22.70 | 4.76 | 22.38 | 4.73 | 21.82 | 4.67 | 22.81 | 4.78 |
| Model 4 | School ID | 0.06 | 0.23 | 0.06 | 0.25 | 0.06 | 0.24 | 0.06 | 0.25 | 0.06 | 0.24 |
|  | Residual | 19.20 | 4.38 | 20.27 | 4.50 | 20.00 | 4.47 | 19.93 | 4.46 | 20.27 | 4.50 |

Note: Model 1: unadjusted model; Model 2: adjusted for demographic variables; Model 3: further adjusted for behavioural variables; Model 4: further adjusted for social variables. SD: standard deviation

Table S4. Within- and between-group variability for the association between SDQTD score and SWEMWBS for 114,439 individuals across 193 schools (complete case analysis)

|  |  | Variance | SD |
| --- | --- | --- | --- |
| Model 1 | School ID | 0.07 | 0.26 |
|  | Residual | 20.20 | 4.49 |
| Model 2 | School ID | 0.06 | 0.25 |
|  | Residual | 19.52 | 4.42 |
| Model 3 | School ID | 0.05 | 0.23 |
|  | Residual | 17.92 | 4.23 |
| Model 4 | School ID | 0.05 | 0.22 |
|  | Residual | 16.52 | 4.07 |

Note: Model 1: unadjusted model; Model 2: adjusted for demographic variables; Model 3: further adjusted for behavioural variables; Model 4: further adjusted for social variables. SD: standard deviation

*Univariable and multivariable multilevel linear regression – complete case analysis (N=114,439)*

Table S5 presents the association between SDQ total score and SWEMWBS using univariable and multivariable multilevel linear regression for the complete case analysis. The intercept was 26.11, indicating that those registering as “Close to average” on SDQTD experience moderate wellbeing (defined as 21-27 on the SWEWMBS scale). There was evidence of a clear dose-response relationship indicated by increasing severity of difficulties accompanied by a marked drop in wellbeing scores across all models (Models 1 to 4). For example, in the unadjusted model, individuals scoring “slightly raised” on the SDQTD were associated with lower SWEMSWBS scores (*b* = -3.47, 95% CI = -3.55 to -3.39, p < 2e-16) compared to individuals scoring “close to average” on the SDQTD. Individuals scoring “high” were associated with lower SWEMSWBS scores (*b* = -4.78, 95% CI = -4.88 to -4.67, p < 2e-16) compared to individuals scoring “close to average”. Finally, individuals scoring “very high” on the SDQTD were associated with lower SWEMWBS scores (*b* = -7.13, 95% CI = -7.20 to -7.06, p < 2e-16) compared to individuals who scored “close to average” on the SDQTD. Focusing on clinical implications, individuals in the “very high” group resulted in a SWEMWBS score of 19, which is near the lower bound of possible mild depression according to the categorical interpretation of SWEMWBS scores (Warwick Medical School, 2021).

The same pattern of results was observed for all additional models (Models 2-4). Focusing on the fully adjusted models (Model 4: adjusting for demographic, behavioural, and social domains), estimates attenuated a little, however still remained statically significant. For example, individuals scoring “slightly raised” on the SDQTD were associated with lower SWEMSWBS scores (*b* = -1.40, 95% CI = -1.46 to -1.34, p < 2e-16) compared to individuals scoring “close to average” on the SDQTD. Individuals scoring “high” were associated with lower SWEMSWBS scores (*b* = -2.03, 95% CI = -2.11 to -1.95, p < 2e-16) compared to individuals scoring “close to average”. Finally, individuals scoring “very high” on the SDQTD were associated with lower SWEMWBS scores (*b* = -3.14, 95% CI = -3.20 to -3.08, p < 2e-16) compared to individuals who scored “close to average” on the SDQTD. The intercept was 24.03, indicating that those registering as “Close to average” on SDQTD experience moderate wellbeing (defined as 21-27 on the SWEWMBS scale). Focusing on clinical implications, individuals in the “very high” group resulted in a SWEMWBS score of 21, which is near the lower bound of possible mild depression according to the categorical interpretation of SWEMWBS scores (Warwick Medical School, 2021c).

*Sensitivity analyses*

Results from the complete case analyses examining the association between SDQTD score and SWEMWBS showed a similar pattern to our primary results using multiple imputed data (Supplementary Material Table S5). The association between SDQTD and SWEMWBS in the complete case analysis produced slightly larger estimates across all SDQTD categories and across all models. Focusing on the fully adjusted model (Model 4), individuals scoring ‘Slightly raised’ were associated with lower SWEMWBS scores (*b* = -1.68, 95% CI = -1.76 to -1.61, p<2e-16) compared to individuals scoring ‘Close to average’; individuals scoring ‘High’ were associated with lower SWEMWBS scores (*b* = -2.41, 95% CI = -2.51 to -2.31, p<2e-16) compared to individuals scoring ‘Close to average’. Finally, individuals scoring ‘Very high’ were associated with lower SWEMWBS scores (*b* = -3.63, 95% CI = -3.71 to -3.55, p<2e-16) compared to individuals who scored ‘Close to average’. Results from the complete case analyses examining the association between each of the SDQ subscales and SWEMWBS showed a similar pattern to our primary results using multiple imputed data (Supplementary Material Table S6).

Results for the multiple imputation sample (N=212,975) examining the association between SDQ Total Difficulties and SWEMWBS (Supplementary Material Table S7) and SDQ subscales and SWEMWBS (Supplementary Material Table S8) showed a similar pattern of results to the primary analyses. Results for the multiple imputation sample for assessment year 2019/20 (N=105,934) examining the association between SDQ Total Difficulties and SWEMWBS (Supplementary Material Table S9) and SDQ subscales and SWEMWBS (Supplementary Material Table S10) showed a similar pattern of results to the primary analyses. Results for the multiple imputation sample for assessment year 2021/22 (N=109,031) examining the association between SDQ Total Difficulties and SWEMWBS (Supplementary Material Table S11) and SDQ subscales and SWEMWBS (Supplementary Material Table S12) also showed a similar pattern of results to the primary analyses.

Table S5. Association between SDQTD score and SWEMWBS for *114,439* individuals aged 11-16 in Welsh Secondary schools from 2019-2021. Intercept, Estimates and their 95% confidence intervals are included for each of the 4 models.

|  | Model 1 | | Model 2 | | Model 3 | | Model 4 | |
| --- | --- | --- | --- | --- | --- | --- | --- | --- |
|  | *b*  (95%CI) | *p* | *b*  (95%CI) | *p* | *b*  (95%CI) | *p* | *b*  (95%CI) | *p* |
| *Intercept* | 26.11  (26.04, 26.19) | <2e-16 | 27.07  (26.97, 27.17) | <2e-16 | 24.42  (24.04, 24.80) | <2e-16 | 25.43  (25.06, 25.80) | <2e-16 |
| *SDQ (close to average)* | Reference group | | Reference group | | Reference group | | Reference group | |
| *SDQ 15-17 (Slightly raised)* | -3.47  (-3.55, -3.39) | <2e-16 | -3.23  (-3.30, -3.15) | <2e-16 | -2.26  (-2.34, -2.18) | <2e-16 | -1.68  (-1.76, -1.61) | <2e-16 |
| *SDQ 18-19 (High)* | -4.78  (-4.88, -4.67) | <2e-16 | -4.46  (-4.56, -4.36) | <2e-16 | -3.18  (-3.28, -3.08) | <2e-16 | -2.41  (-2.51, -2.31) | <2e-16 |
| *SDQ 20-40 (Very high)* | -7.12  (-7.20, -7.06) | <2e-16 | -6.65  (-6.72, -6.58) | <2e-16 | -4.84  (-4.92, -4.76) | <2e-16 | -3.63  (-3.71, -3.55) | <2e-16 |

Note: Model 1: unadjusted model; Model 2: adjusted for demographic variables; Model 3: further adjusted for behavioural variables; Model 4: further adjusted for social variables.

*Univariable and multivariable multilevel linear regression – complete case analysis*

Table S6 presents the association between the SDQ sub scales and SWEMWBS using univariable and multivariable multilevel regression for the complete case analysis. There was evidence of a clear dose-response relationship indicated by increasing severity of all SDQ subscales accompanied by a marked drop in wellbeing scores across all models (Models 1 to 4). The strongest association was between the emotional difficulties subscale and SWEMWBS. Focusing on the fully adjusted model, the intercept was 25.08, indicating that those registering as “Close to average” on SDQ emotional difficulties subscale experience moderate wellbeing (defined as 21-27 on the SWEWMBS scale). For example, in the fully adjusted model, individuals scoring “slightly raised” on the SDQ emotional difficulties subscale were associated with lower SWEMSWBS scores (*b* = -1.52, 95% CI = -1.60 to -1.44, p<2e-16) compared to individuals scoring “close to average” on the SDQTD. Individuals scoring “high” were associated with lower SWEMSWBS scores (*b* = -2.07, 95% CI = -2.15 to -1.98, p<2e-16) compared to individuals scoring “close to average”. Finally, individuals scoring “very high” on the SDQ emotional difficulties subscale were associated with lower SWEMWBS scores (*b* = -3.35, 95% CI = -3.43 to -3.28, p<2e-16) compared to individuals who scored “close to average” on the SDQ emotional difficulties subscale.

Table S6. Association between SDQ subscales and SWEMWBS for *114,439* individuals aged 11-16 in Welsh Secondary schools from 2019-2021. Intercept, Estimates and their 95% confidence intervals are included for each of the 4 models.

| Emotional difficulties | | | | | | | | |
| --- | --- | --- | --- | --- | --- | --- | --- | --- |
|  | Model 1 | | Model 2 | | Model 3 | | Model 4 | |
|  | *b*  (95%CI) | *p* | *b*  (95%CI) | *p* | *b*  (95%CI) | *p* | *b*  (95%CI) | *p* |
| *Intercept* | 26.10  (26.00, 26.18) | <2e-16 | 26.70  (26.62, 26.84) | <2e-16 | 23.63  (23.25, 24.02) | <2e-16 | 25.08  (24.71, 25.44) | <2e-16 |
| *SDQ (0-4)*  *Close to average* | Reference group | | Reference group | | Reference group | | Reference group | |
| *SDQ (5)*  *Slightly raised* | -3.06  (-3.16, -2.97) | <2e-16 | -2.87  (-2.96, -2.78) | <2e-16 | -1.97  (-2.06, -1.89) | <2e-16 | -1.52  (-1.60, -1.44) | <2e-16 |
| *SDQ (6)*  *High* | -4.12  (-4.22, -4.03) | <2e-16 | -3.85  (-3.95, -3.76) | <2e-16 | -2.67  (-2.76, -2.58) | <2e-16 | -2.07  (-2.15, -1.98) | <2e-16 |
| *SDQ (7-10)*  *Very high* | -6.61  (-6.67, -6.54) | <2e-16 | -6.15  (-6.22, -6.08) | <2e-16 | -4.33  (-4.40, -4.26) | <2e-16 | -3.35  (-3.43, -3.28) | <2e-16 |
| Conduct problems | | | | | | | | |
|  | Model 1 | | Model 2 | | Model 3 | | Model 4 | |
|  | *b*  (95%CI) | *p* | *b*  (95%CI) | *p* | *b*  (95%CI) | *p* | *b*  (95%CI) | *P* |
| *Intercept* | 24.80  (24.72, 24.90) | <2e-16 | 26.60  (26.48, 26.73) | <2e-16 | 23.97  (23.57, 24.37) | <2e-16 | 25.42  (25.04, 25.80) | <2e-16 |
| *SDQ (0-3)*  *Close to average* | Reference group | | Reference group | | Reference group | | Reference group | |
| *SDQ (4)*  *Slightly raised* | -2.67  (-2.77, -2.60) | <2e-16 | -2.61  (-2.70, -2.51) | <2e-16 | -1.44  (-1.53, -1.35) | <2e-16 | -0.85  (-0.93, -0.76) | <2e-16 |
| *SDQ (5)*  *High* | -3.57  (-3.69, -3.50) | <2e-16 | -3.46  (-3.58, -3.34) | <2e-16 | -1.98  (-2.09, -1.88) | <2e-16 | -1.22  (-1.32, -1.11) | <2e-16 |
| *SDQ (6-10)*  *Very high* | -4.66  (-4.78, -4.54) | <2e-16 | -4.41  (-4.53, -4.300 | <2e-16 | -2.34  (-2.45, -2.22) | <2e-16 | -1.38  (-1.49, -1.27) | <2e-16 |
| Hyperactivity | | | | | | | | |
|  | Model 1 | | Model 2 | | Model 3 | | Model 4 | |
|  | *b*  (95%CI) | *p* | *b*  (95%CI) | *P* | *b*  (95%CI) | *p* | *b*  (95%CI) | *p* |
| *Intercept* | 25.41  (25.29, 25.48) | <2e-16 | 26.85  (26.73, 26.96) | <2e-16 | 23.72  (23.30, 24.12) | <2e-16 | 25.32  (24.94, 25.69) | <2e-16 |
| *SDQ (0-5)*  *Close to average* | Reference group | | Reference group | | Reference group | | Reference group | |
| *SDQ (6)*  *Slightly raised* | -2.55  (-2.65, -2.46) | <2e-16 | -2.41  (-2.49, -2.32) | <2e-16 | -1.36  (-1.45, -1.28) | <2e-16 | -0.97  (-1.05, -0.89) | <2e-16 |
| *SDQ (7)*  *High* | -3.35  (-3.45, -3.25) | <2e-16 | -3.08  (-3.18, -2.98) | <2e-16 | -1.77  (-1.86, -1.68) | <2e-16 | -1.23  (-1.32, -1.14) | <2e-16 |
| *SDQ (8-10)*  *Very high* | -5.02  (-5.10, -4.94) | <2e-16 | -4.59  (-4.67, -4.51) | <2e-16 | -2.68  (-2.77, -2.61) | <2e-16 | -1.87  (-1.95, -1.80) | <2e-16 |
| Peer problems | | | | | | | | |
|  | Model 1 | | Model 2 | | Model 3 | | Model 4 | |
|  | *b*  (95%CI) | *p* | *b*  (95%CI) | *P* | *b*  (95%CI) | *p* | *b*  (95%CI) | *P* |
| *Intercept* | 25.31  (25.23, 25.42) | <2e-16 | 27.02  (26.90, 27.14) | <2e-16 | 24.71  (24.31, 25.11) | <2e-16 | 25.71  (25.33, 26.09) | <2e-16 |
| *SDQ (0-2)*  *Close to average* | Reference group | | Reference group | | Reference group | | Reference group | |
| *SDQ (3)*  *(Slightly raised* | -2.54  (-2.62, -2.45) | <2e-16 | -2.39  (-2.47, -2.31) | <2e-16 | -1.55  (-1.63, -1.48) | <2e-16 | -1.06  (-1.14, -0.99) | <2e-16 |
| *SDQ (4)*  *High* | -3.77  (-3.87, -3.67) | <2e-16 | -3.58  (-3.68, -3.48) | <2e-16 | -2.38  (-2.47, -2.30) | <2e-16 | -1.55  (-1.64, -1.47) | <2e-16 |
| *SDQ (5-10)*  *Very high* | -5.48  (-5.58, -5.39) | <2e-16 | -5.20  (-5.30, -5.11) | <2e-16 | -3.57  (-3.66, -3.48) | <2e-16 | -2.32  (-2.41, -2.23) | <2e-16 |
| Prosocial | | | | | | | | |
|  | Model 1 | | Model 2 | | Model 3 | | Model 4 | |
|  | *b*  (95%CI) | *p* | *b*  (95%CI) | *P* | *b*  (95%CI) | *P* | *b*  (95%CI) | *P* |
| *Intercept* | 24.61  (24.26, 24.67) | <2e-16 | 26.60  (26.46, 26.72) | <2e-16 | 24.06  (23.65, 24.46) | <2e-16 | 25.58  (25.20, 25.97) | <2e-16 |
| *SDQ (7-10)*  *Close to average* | Reference group | | Reference group | | Reference group | | Reference group | |
| *SDQ (6)*  *Slightly raised* | -1.23  (-1.32, -1.14) | <2e-16 | -1.50  (-1.59, -1.41) | <2e-16 | -1.01  (-1.09, -0.93) | <2e-16 | -0.73  (-0.80, -0.65) | <2e-16 |
| *SDQ (5)*  *High* | -1.69  (-1.80, -1.59) | <2e-16 | -2.02  (-2.12, -1.92) | <2e-16 | -1.28  (-1.37, -1.19) | <2e-16 | -0.87  (-0.96, -0.78) | <2e-16 |
| *SDQ (0-4)*  *Very high* | -2.68  (-2.79, -2.58) | <2e-16 | -3.01  (-3.11, -2.90) | <2e-16 | -1.78  (-1.87, -1.68) | <2e-16 | -1.10  (-1.20, -1.01) | <2e-16 |

Note: Model 1: unadjusted model; Model 2: adjusted for demographic variables; Model 3: further adjusted for behavioural variables; Model 4: further adjusted for social variables. P values are adjusted for Bonferroni correction.

*Univariable and multivariable multilevel linear regression – Multiple imputation (N=212,975)*

Table S7 presents the association between SDQ total score and SWEMWBS using univariable and multivariable multilevel linear regression for 212,975 individuals. The intercept was 25.44, indicating that those registering as “Close to average” on SDQTD experience moderate wellbeing (defined as 21-27 on the SWEWMBS scale). There was evidence of a clear dose-response relationship indicated by increasing severity of difficulties accompanied by a marked drop in wellbeing scores across all models (Models 1 to 4). For example, in the unadjusted model, individuals scoring “slightly raised” on the SDQTD were associated with lower SWEMSWBS scores (*b* = -3.18, 95% CI = -3.53 to -2.83, p < 4e-16) compared to individuals scoring “close to average” on the SDQTD. Individuals scoring “high” were associated with lower SWEMSWBS scores (*b* = -4.26, 95% CI = -4.56 to -3.96, p < 2e-16) compared to individuals scoring “close to average”. Finally, individuals scoring “very high” on the SDQTD were associated with lower SWEMWBS scores (*b* = -6.32, 95% CI = -6.83 to -5.81, p < 2e-16) compared to individuals who scored “close to average” on the SDQTD. Focusing on clinical implications, individuals in the “very high” group resulted in a SWEMWBS score of 19, which is near the lower bound of possible mild depression according to the categorical interpretation of SWEMWBS scores (Warwick Medical School, 2021c).

The same pattern of results was observed for all additional models (Models 2-4). Focusing on the fully adjusted models (Model 4: adjusting for demographic, behavioural, and social domains), estimates attenuated a little, however still remained statically significant. For example, individuals scoring “slightly raised” on the SDQTD were associated with lower SWEMSWBS scores (*b* = -1.55, 95% CI = -1.75 to -1.34, p < 6e-15) compared to individuals scoring “close to average” on the SDQTD. Individuals scoring “high” were associated with lower SWEMSWBS scores (*b* = -2.12, 95% CI = -2.29 to -1.96, p < 2e-16) compared to individuals scoring “close to average”. Finally, individuals scoring “very high” on the SDQTD were associated with lower SWEMWBS scores (*b* = -3.14, 95% CI = -3.37 to -2.92, p < 2e-16) compared to individuals who scored “close to average” on the SDQTD. The intercept was 24.03, indicating that those registering as “Close to average” on SDQTD experience moderate wellbeing (defined as 21-27 on the SWEWMBS scale). Focusing on clinical implications, individuals in the “very high” group resulted in a SWEMWBS score of 21, which is near the lower bound of possible mild depression according to the categorical interpretation of SWEMWBS scores (Warwick Medical School, 2021c).

Table S7. Association between SDQTD score and SWEMWBS for 212,951 individuals aged 11-16 in Welsh Secondary schools from 2019-2021. Intercept, Estimates and their 95% confidence intervals are included for each of the 4 models.

|  | Model 1 | | Model 2 | | Model 3 | | Model 4 | |
| --- | --- | --- | --- | --- | --- | --- | --- | --- |
|  | *b*  (95%CI) | *p* | *b*  (95%CI) | *p* | *b*  (95%CI) | *p* | *b*  (95%CI) | *p* |
| *Intercept* | 25.44  (25.10, 25.79) | <2e-16 | 26.49  (25.96, 27.03) | <2e-16 | 25.49  (24.85, 26.14) | <2e-16 | 26.30  (25.95, 26.66) | <2e-15 |
| *SDQ*  *(Close to average)* | Reference group | | Reference group | | Reference group | | Reference group | |
| *SDQ 15-17*  *(Slightly raised)* | -3.18  (-3.53, -2.83) | <4e-16 | -2.96  (-3.30, -2.61) | <2e-15 | -2.06  (-2.36, -1.86) | <3e-13 | -1.55  (-1.75, -1.34) | <6e-15 |
| *SDQ 18-19 (High)* | -4.26  (-4.56, -3.96) | <2e-16 | -3.98  (-4.29, -3.67) | <2e-16 | -2.83  (-3.11, -2.55) | <2e-16 | -2.12  (-2.29, -1.96) | <2e-16 |
| *SDQ 20-40*  *(Very high)* | -6.32  (-6.83, -5.81) | <2e-16 | -5.91  (-6.43, -5.39) | <2e-16 | -4.35  (-4.81, -3.89) | <5e-16 | -3.14  (-3.37, -2.92) | <2e-16 |

Note: Model 1: unadjusted model; Model 2: adjusted for demographic variables; Model 3: further adjusted for behavioural variables; Model 4: further adjusted for social variables.

*Univariable and multivariable multilevel linear regression – Multiple imputation (N=212,975)*

Table S8 presents the association between the SDQ sub scales and SWEMWBS using univariable and multivariable multilevel regression for 212,975 individuals. There was evidence of a clear dose-response relationship indicated by increasing severity of all SDQ subscales accompanied by a marked drop in wellbeing scores across all models (Models 1 to 4). The strongest association was between the emotional difficulties subscale and SWEMWBS. Focusing on the fully adjusted model, the intercept was 25.08, indicating that those registering as “Close to average” on SDQ emotional difficulties subscale experience moderate wellbeing (defined as 21-27 on the SWEWMBS scale). For example, in the fully adjusted model, individuals scoring “slightly raised” on the SDQ emotional difficulties subscale were associated with lower SWEMSWBS scores (*b* = -1.52, 95% CI = -1.63 to -1.40, p<2e-16) compared to individuals scoring “close to average” on the SDQ emotional difficulties subscale. Individuals scoring “high” were associated with lower SWEMSWBS scores (*b* = -1.99, 95% CI = -2.14 to -1.86, p<2e-16) compared to individuals scoring “close to average”. Finally, individuals scoring “very high” on the SDQ emotional difficulties subscale were associated with lower SWEMWBS scores (*b* = -3.25, 95% CI = -3.42 to -3.08, p<2e-16) compared to individuals who scored “close to average” on the SDQ emotional difficulties subscale.

Table S8. Association between SDQ subscales and SWEMWBS for 212,975 individuals aged 11-16 in Welsh Secondary schools from 2019-2021. Intercept, Estimates and their 95% confidence intervals are included for each of the 4 models.

| Emotional difficulties | | | | | | | | | | | | |
| --- | --- | --- | --- | --- | --- | --- | --- | --- | --- | --- | --- | --- |
|  | Model 1 | | | Model 2 | | | Model 3 | | | Model 4 | | |
|  | *b*  (95%CI) | *p* | | *b*  (95%CI) | | *p* | *b*  (95%CI) | | *p* | *b*  (95%CI) | | *p* |
| *Intercept* | 25.37  (25.00, 25.73) | <2e-16 | | 26.06  (25.51, 26.62) | | <2e-16 | 25.21  (24.55, 25.88) | | <2e-16 | 26.24  (25.89, 26.61) | | <2e-16 |
| *SDQ (0-4)*  *Close to average* | Reference group | | | Reference group | | | Reference group | | | Reference group | | |
| *SDQ (5)*  *Slightly raised* | -2.90  (-3.07, -2.73) | <2e-16 | | -2.73  (-2.91, -2.56) | | <2e-16 | -1.90  (-2.06, -1.74) | | <2e-16 | -1.52  (-1.63, -1.40) | | <2e-16 |
| *SDQ (6)*  *High* | -3.83  (-4.08, -3.56) | <2e-16 | | -3.57  (-3.83, -3.31) | | <2e-16 | -2.49  (-2.72, -2.27) | | <2e-16 | -1.99  (-2.14, -1.86) | | <2e-16 |
| *SDQ (7-10)*  *Very high* | -6.16  (-6.50, -5.81) | <2e-16 | | -5.74  (-6.11, -5.36) | | <2e-16 | -4.15  (-4.46, -3.85) | | <2e-16 | -3.25  (-3.42, -3.08) | | <2e-16 |
| Conduct problems | | | | | | | | | | | | |
|  | Model 1 | | | Model 2 | | | Model 3 | | | Model 4 | | |
|  | *b*  (95%CI) | | *p* | *b*  (95%CI) | *p* | | *b*  (95%CI) | *p* | | *b*  (95%CI) | *P* | |
| *Intercept* | 24.06  (23.69, 24.42) | | <2e-16 | 25.96  (25.42, 26.51) | <2e-16 | | 25.13  (24.48, 25.78) | <2e-16 | | 26.25  (25.89, 26.62) | <2e-16 | |
| *SDQ (0-3)*  *Close to average* | Reference group | | | Reference group | | | Reference group | | | Reference group | | |
| *SDQ (4)*  *Slightly raised* | -2.49  (-2.74, -2.23) | | <2e-16 | -2.45  (-2.70, -2.20) | <2e-16 | | -1.56  (-1.77, -1.34) | <2e-16 | | -0.90  (-1.02, -0.77) | <2e-16 | |
| *SDQ (5)*  *High* | -3.11  (-3.46, -2.75) | | <2e-16 | -3.08  (-3.44, -2.72) | <2e-16 | | -2.01  (-2.32, -1.71) | <2e-15 | | -1.15  (-1.32, -0.99) | <2e-16 | |
| *SDQ (6-10)*  *Very high* | -3.86  (-4.43, -3.29) | | <8e-13 | -3.77  (-4.33, -3.20) | <1e-12 | | -2.36  (-2.82, -1.90) | <2e-13 | | -1.26  (-1.49, -1.04) | <2e-12 | |
| Hyperactivity | | | | | | | | | | | | |
|  | Model 1 | | | Model 2 | | | Model 3 | | | Model 4 | | |
|  | *b*  (95%CI) | | *p* | *b*  (95%CI) | *P* | | *b*  (95%CI) | *p* | | *b*  (95%CI) | *p* | |
| *Intercept* | 24.61  (24.24, 24.99) | | <2e-16 | 26.05  (25.49, 26.61) | <2e-16 | | 25.18  (24.51, 25.84) | <2e-16 | | 26.32  (25.95, 26.68) | <2e-16 | |
| *SDQ (0-5)*  *Close to average* | Reference group | | | Reference group | | | Reference group | | | Reference group | | |
| *SDQ (6)*  *Slightly raised* | -2.47  (-2.71, -2.23) | | <2e-16 | -2.34  (-2.57, -2.10) | <2e-16 | | -1.46  (-1.65, -1.27) | <2e-16 | | -1.04  (-1.16, -0.92) | <2e-16 | |
| *SDQ (7)*  *High* | -3.08  (-3.31, -2.85) | | <2e-16 | -2.84  (-3.07, -2.60) | <2e-16 | | -1.77  (-1.96, -1.58) | <2e-16 | | -1.23  (-1.38, -1.09) | <2e-16 | |
| *SDQ (8-10)*  *Very high* | -4.52  (-4.91, -4.13) | | <2e-16 | -4.14  (-4.53, -3.75) | <2e-16 | | -2.69  (-3.01, -2.37) | <2e-15 | | -1.89  (-2.02, -1.66) | <2e-16 | |
| Peer problems | | | | | | | | | | | | |
|  | Model 1 | | | Model 2 | | | Model 3 | | | Model 4 | | |
|  | *b*  (95%CI) | | *p* | *b*  (95%CI) | *P* | | *b*  (95%CI) | *p* | | *b*  (95%CI) | *P* | |
| *Intercept* | 24.68  (24.34, 25.03) | | <2e-16 | 26.48  (25.97, 26.99) | <2e-16 | | 25.75  (25.14, 26.35) | <2e-16 | | 26.51  (26.16, 26.87) | <2e-16 | |
| *SDQ (0-2)*  *Close to average* | Reference group | | | Reference group | | | Reference group | | | Reference group | | |
| *SDQ (3)*  *(Slightly raised* | -2.50  (-2.73, -2.26) | | <2e-16 | -2.36  (-2.59, -2.12) | <2e-16 | | -1.61  (-1.81, -1.40) | <2e-15 | | -1.12  (-1.28, -0.97) | <2e-16 | |
| *SDQ (4)*  *High* | -3.74  (-4.14, -3.35) | | <2e-16 | -3.61  (-4.01, -3.21) | <2e-16 | | -2.55  (-2.90, -2.21) | <4e-14 | | -1.70  (-1.90, -1.49) | <2e-16 | |
| *SDQ (5-10)*  *Very high* | -5.02  (-5.60, -4.43) | | <6e-15 | -4.85  (-5.43, -4.26) | <2e-16 | | -3.48  (-4.00, -2.95) | <2e-12 | | -2.20  (-2.46, -1.93) | <2e-16 | |
| Prosocial behaviour | | | | | | | | | | | | |
|  | Model 1 | | | Model 2 | | | Model 3 | | | Model 4 | | |
|  | *b*  (95%CI) | | *p* | *b*  (95%CI) | *P* | | *b*  (95%CI) | *P* | | *b*  (95%CI) | *P* | |
| *Intercept* | 23.84  (23.44, 24.23) | | <2e-16 | 25.93  (25.37, 26.49) | <2e-16 | | 25.54  (24.90, 26.17) | <2e-16 | | 26.55  (26.21, 26.89) | <2e-16 | |
| *SDQ (7-10)*  *Close to average* | Reference group | | | Reference group | | | Reference group | | | Reference group | | |
| *SDQ (6)*  *Slightly raised* | -1.25  (-1.44, -1.06) | | <2e-14 | -1.52  (-1.71, -1.32) | <2e-16 | | -1.16  (-1.34, -0.99) | <8e-15 | | -0.78  (-0.94, -0.63) | <5e-12 | |
| *SDQ (5)*  *High* | -1.78  (-2.06, -1.50) | | <3e-13 | -2.12  (-2.41, -1.82) | <2e-14 | | -1.59  (-1.86, -1.33) | <2e-12 | | -1.00  (-1.23, -0.77) | <2e-09 | |
| *SDQ (0-4)*  *Very high* | -2.92  (-3.54, -2.31) | | <2e-09 | -3.31  (-3.94, -2.67) | <2e-10 | | -2.45  (-3.00, -1.90) | <5e-09 | | -1.47  (-1.85, -1.09) | <5e-08 | |

Note: Model 1: unadjusted model; Model 2: adjusted for demographic variables; Model 3: further adjusted for behavioural variables; Model 4: further adjusted for social variables. P values are adjusted for Bonferroni correction.

*Univariable and multivariable multilevel linear regression for 2019/20 survey – Multiple imputation (N=105,934)*

Table S9 presents the association between the SDQTD and SWEMWBS using univariable and multivariable multilevel regression for the 2019/20 survey including 105,934 individuals. There was evidence of a clear dose-response relationship indicated by increasing severity of the SDQTD accompanied by a marked drop in wellbeing scores across all models (Models 1 to 4). Focusing on the fully adjusted model (Model 4), the intercept was 26.42, indicating that those registering as “Close to average” on SDQTD experience moderate wellbeing (defined as 21-27 on the SWEWMBS scale). For example, in the fully adjusted model, individuals scoring “slightly raised” on the SDQTD were associated with lower SWEMSWBS scores (*b* = -1.61, 95% CI = -1.71 to -1.50, p<2e-16) compared to individuals scoring “close to average” on the SDQTD. Individuals scoring “high” were associated with lower SWEMSWBS scores (*b* = -2.02, 95% CI = -2.13 to -1.90, p<2e-16) compared to individuals scoring “close to average” on the SDQTD. Finally, individuals scoring “very high” on the SDQTD were associated with lower SWEMWBS scores (*b* = -3.50, 95% CI = -3.59 to -3.40, p<2e-16) compared to individuals who scored “close to average” on the SDQTD. Results for the association between the SDQ sub scales and SWEMWBS are presented in Table S9.

Table S9. Association between SDQTD score and SWEMWBS for 105,934 individuals aged 11-16 in Welsh Secondary schools from 2019-2021. Intercept, Estimates and their 95% confidence intervals are included for each of the 4 models (2019 survey).

|  | Model 1 | | Model 2 | | Model 3 | | Model 4 | |
| --- | --- | --- | --- | --- | --- | --- | --- | --- |
|  | *b*  (95%CI) | *p* | *b*  (95%CI) | *p* | *b*  (95%CI) | *p* | *b*  (95%CI) | *p* |
| *Intercept* | 25.75  (25.69, 25.82) | <2e-16 | 26.61  (26.53, 26.69) | <2e-16 | 25.68  (25.52, 25.85) | <2e-16 | 26.51  (26.67, 26.67) | <2e-16 |
| *SDQ*  *(Close to average)* | Reference group | | Reference group | | Reference group | | Reference group | |
| *SDQ 15-17*  *(Slightly raised)* | -3.26  (-3.36, -3.15) | <2e-16 | -3.10  (-3.21, -3.00) | <2e-16 | -2.26  (-2.35, -2.16) | <2e-16 | -1.73  (-1.83, -1.64) | <2e-16 |
| *SDQ 18-19 (High)* | -4.59  (-4.72, -4.45) | <2e-16 | -4.38  (-4.52, -4.25) | <2e-16 | -3.21  (-3.34, -3.08) | <2e-16 | -2.46  (-2.59, -2.33) | <2e-16 |
| *SDQ 20-40*  *(Very high)* | -6.80  (-6.89, -6.72) | <2e-16 | -6.52  (-6.61, -6.43) | <2e-16 | -4.92  (-5.01, -4.83) | <2e-16 | -3.71  (-3.81, -3.61) | <2e-16 |

Note: Model 1: unadjusted model; Model 2: adjusted for demographic variables; Model 3: further adjusted for behavioural variables; Model 4: further adjusted for social variables.

Table S10. Association between SDQ subscales and SWEMWBS for 105,934 individuals aged 11-16 in Welsh Secondary schools from 2019-2021. Intercept, Estimates and their 95% confidence intervals are included for each of the 4 models (2019 survey).

| Emotional difficulties | | | | | | | | | | | | |
| --- | --- | --- | --- | --- | --- | --- | --- | --- | --- | --- | --- | --- |
|  | Model 1 | | | Model 2 | | | Model 3 | | | Model 4 | | |
|  | *b*  (95%CI) | *p* | | *b*  (95%CI) | | *p* | *b*  (95%CI) | | *p* | *b*  (95%CI) | | *p* |
| *Intercept* | 25.69  (25.61, 25.76) | <2e-16 | | 26.13  (26.05, 26.22) | | <2e-16 | 25.34  (25.18, 25.50) | | <2e-16 | 26.42  (26.25, 26.59) | | <2e-16 |
| *SDQ (0-4)*  *Close to average* | Reference group | | | Reference group | | | Reference group | | | Reference group | | |
| *SDQ (5)*  *Slightly raised* | -2.91  (-3.02, -2.80) | <2e-16 | | -2.79  (-2.91, -2.68) | | <2e-16 | -1.99  (-2.09, -1.88) | | <2e-16 | -1.61  (-1.71, -1.50) | | <2e-16 |
| *SDQ (6)*  *High* | -3.75  (-3.87, -3.62) | <2e-16 | | -3.59  (-3.71, -3.47) | | <2e-16 | -2.52  (-2.63, -2.41) | | <2e-16 | -2.02  (-2.13, -1.90) | | <2e-16 |
| *SDQ (7-10)*  *Very high* | -6.35  (-6.44, -6.26) | <2e-16 | | -6.08  (-6.17, -5.99) | | <2e-16 | -4.43  (-4.52, -4.33) | | <2e-16 | -3.50  (-3.59, -3.40) | | <2e-16 |
| Conduct problems | | | | | | | | | | | | |
|  | Model 1 | | | Model 2 | | | Model 3 | | | Model 4 | | |
|  | *b*  (95%CI) | | *p* | *b*  (95%CI) | *p* | | *b*  (95%CI) | *p* | | *b*  (95%CI) | *P* | |
| *Intercept* | 24.57  (24.49, 24.65) | | <2e-16 | 26.12  (26.02, 26.21) | <2e-16 | | 25.26  (25.10, 25.43) | <2e-16 | | 26.42  (26.26, 26.59) | <2e-16 | |
| *SDQ (0-3)*  *Close to average* | Reference group | | | Reference group | | | Reference group | | | Reference group | | |
| *SDQ (4)*  *Slightly raised* | -2.50  (-2.62, -2.39) | | <2e-16 | -2.51  (-2.63, -2.39) | <2e-16 | | -1.65  (-1.76, -1.53) | <2e-16 | | -0.99  (-1.10, -0.89) | <2e-16 | |
| *SDQ (5)*  *High* | -3.33  (-3.46, -3.19) | | <2e-16 | -3.36  (-3.50, -3.23) | <2e-16 | | -2.25  (-2.38, -2.11) | <2e-16 | | -1.37  (-1.50, -1.25) | <2e-16 | |
| *SDQ (6-10)*  *Very high* | -4.30  (-4.44, -4.17) | | <2e-16 | -4.29  (-4.42, -4.16) | <2e-16 | | -2.76  (-2.88, -2.63) | <2e-16 | | -1.64  (-1.77, -1.52) | <2e-16 | |
| Hyperactivity | | | | | | | | | | | | |
|  | Model 1 | | | Model 2 | | | Model 3 | | | Model 4 | | |
|  | *b*  (95%CI) | | *p* | *b*  (95%CI) | *P* | | *b*  (95%CI) | *p* | | *b*  (95%CI) | *p* | |
| *Intercept* | 24.90  (24.82, 24.98) | | <2e-16 | 26.22  (26.12, 26.30) | <2e-16 | | 25.35  (25.18, 25.52) | <2e-16 | | 26.53  (26.36, 26.70) | <2e-16 | |
| *SDQ (0-5)*  *Close to average* | Reference group | | | Reference group | | | Reference group | | | Reference group | | |
| *SDQ (6)*  *Slightly raised* | -2.42  (-2.53, -2.30) | | <2e-16 | -2.36  (-2.47, -2.25) | <2e-16 | | -1.48  (-1.59, -1.37) | <2e-16 | | -1.07  (-1.17, -0.97) | <2e-16 | |
| *SDQ (7)*  *High* | -3.04  (-3.18, -2.91) | | <2e-16 | -2.90  (-3.03, -2.77) | <2e-16 | | -1.81  (-1.94, -1.69) | <2e-16 | | -1.30  (-1.42, -1.18) | <2e-16 | |
| *SDQ (8-10)*  *Very high* | -4.57  (-4.68, -4.47) | | <2e-16 | -4.37  (-4.48, -4.27) | <2e-16 | | -2.85  (-2.95, -2.75) | <2e-15 | | -2.00  (-2.09, -1.90) | <2e-16 | |
| Peer problems | | | | | | | | | | | | |
|  | Model 1 | | | Model 2 | | | Model 3 | | | Model 4 | | |
|  | *b*  (95%CI) | | *p* | *b*  (95%CI) | *P* | | *b*  (95%CI) | *p* | | *b*  (95%CI) | *P* | |
| *Intercept* | 25.13  (25.05, 25.21) | | <2e-16 | 26.59  (26.50, 26.68) | <2e-16 | | 25.87  (25.71, 26.04) | <2e-16 | | 26.68  (26.50, 26.83) | <2e-16 | |
| *SDQ (0-2)*  *Close to average* | Reference group | | | Reference group | | | Reference group | | | Reference group | | |
| *SDQ (3)*  *(Slightly raised* | -2.51  (-2.62, -2.41) | | <2e-16 | -2.42  (-2.52, -2.31) | <2e-16 | | -1.67  (-1.76, -1.57) | <2e-16 | | -1.15  (-1.24, -1.05) | <2e-16 | |
| *SDQ (4)*  *High* | -3.83  (-3.96, -3.70) | | <2e-16 | -3.77  (-3.89, -3.64) | <2e-16 | | -2.70  (-2.82, -2.58) | <2e-16 | | -1.85  (-1.96, -1.73) | <2e-16 | |
| *SDQ (5-10)*  *Very high* | -5.51  (-5.63, -5.39) | | <2e-16 | -5.41  (-5.53, -5.29) | <2e-16 | | -3.93  (-4.05, -3.82) | <2e-16 | | -2.59  (-2.72, -2.47) | <2e-16 | |
| Prosocial behaviour | | | | | | | | | | | | |
|  | Model 1 | | | Model 2 | | | Model 3 | | | Model 4 | | |
|  | *b*  (95%CI) | | *p* | *b*  (95%CI) | *P* | | *b*  (95%CI) | *P* | | *b*  (95%CI) | *P* | |
| *Intercept* | 24.32  (23.24, 24.40) | | <2e-16 | 26.07  (25.98, 26.17) | <2e-16 | | 25.63  (25.46, 25.80) | <2e-16 | | 26.71  (26.54, 26.88) | <2e-16 | |
| *SDQ (7-10)*  *Close to average* | Reference group | | | Reference group | | | Reference group | | | Reference group | | |
| *SDQ (6)*  *Slightly raised* | -1.20  (-1.31, -1.09) | | <2e-16 | -1.44  (-1.55, -1.33) | <2e-16 | | -1.10  (-1.20, -1.00) | <2e-16 | | -0.77  (-0.87, -0.78) | <2e-16 | |
| *SDQ (5)*  *High* | -1.77  (-1.91, -1.63) | | <2e-16 | -2.12  (-2.26, -1.98) | <2e-16 | | -1.64  (-1.77, -1.51) | <2e-16 | | -1.07  (-1.19, -0.95) | <2e-16 | |
| *SDQ (0-4)*  *Very high* | -2.78  (-2.90, -2.65) | | <2e-16 | -3.19  (-3.32, -3.07) | <2e-16 | | -2.35  (-2.47, -2.24) | <2e-16 | | -1.45  (-1.56, -1.33) | <2e-16 | |

Note: Model 1: unadjusted model; Model 2: adjusted for demographic variables; Model 3: further adjusted for behavioural variables; Model 4: further adjusted for social variables. P values are adjusted for Bonferroni correction.

*Univariable and multivariable multilevel linear regression for 2021/22 survey – Multiple imputation (N=109,031)*

Table S11 presents the association between the SDQTD and SWEMWBS using univariable and multivariable multilevel regression for the 2021/22 survey including 109,031 individuals. There was evidence of a clear dose-response relationship indicated by increasing severity of the SDQTD accompanied by a marked drop in wellbeing scores across all models (Models 1 to 4). Focusing on the fully adjusted model (Model 4), the intercept was 26.19, indicating that those registering as “Close to average” on SDQTD experience moderate wellbeing (defined as 21-27 on the SWEWMBS scale). For example, in the fully adjusted model, individuals scoring “slightly raised” on the SDQTD were associated with lower SWEMSWBS scores (*b* = -1.63, 95% CI = -1.72 to -1.54, p<2e-16) compared to individuals scoring “close to average” on the SDQTD. Individuals scoring “high” were associated with lower SWEMSWBS scores (*b* = -2.47, 95% CI = -2.58 to -2.36, p<2e-16) compared to individuals scoring “close to average” on the SDQTD. Finally, individuals scoring “very high” on the SDQTD were associated with lower SWEMWBS scores (*b* = -3.67, 95% CI = -3.76 to -3.58, p<2e-16) compared to individuals who scored “close to average” on the SDQTD. Results for the association between the SDQ sub scales and SWEMWBS are presented in Table S11.

Table S11. Association between SDQTD score and SWEMWBS for 109,031 individuals aged 11-16 in Welsh Secondary schools from 2019-2021. Intercept, Estimates and their 95% confidence intervals are included for each of the 4 models (2021 survey).

|  | Model 1 | | Model 2 | | Model 3 | | Model 4 | |
| --- | --- | --- | --- | --- | --- | --- | --- | --- |
|  | *b*  (95%CI) | *p* | *b*  (95%CI) | *p* | *b*  (95%CI) | *p* | *b*  (95%CI) | *p* |
| *Intercept* | 25.52  (25.47, 25.59) | <2e-16 | 26.46  (26.38, 26.53) | <2e-16 | 25.40  (25.25, 25.55) | <2e-16 | 26.19  (26.04, 26.34) | <2e-16 |
| *SDQ*  *(Close to average)* | Reference group | | Reference group | | Reference group | | Reference group | |
| *SDQ 15-17*  *(Slightly raised)* | -3.27  (-3.37, -3.18) | <2e-16 | -3.02  (-3.11, -2.92) | <2e-16 | -2.11  (-2.20, -2.02) | <2e-16 | -1.63  (-1.72, -1.54) | <2e-16 |
| *SDQ 18-19 (High)* | -4.64  (-4.76, -4.52) | <2e-16 | -4.30  (-4.41, -4.19) | <2e-16 | -3.13  (-3.24, -3.02) | <2e-16 | -2.47  (-2.58, -2.36) | <2e-16 |
| *SDQ 20-40*  *(Very high)* | -7.05  (-7.13, -6.98) | <2e-16 | -6.51  (-6.58, -6.43) | <2e-16 | -4.87  (-4.95, -4.78) | <2e-16 | -3.67  (-3.76, -3.58) | <2e-16 |

Note: Model 1: unadjusted model; Model 2: adjusted for demographic variables; Model 3: further adjusted for behavioural variables; Model 4: further adjusted for social variables.

Table S12. Association between SDQ subscales and SWEMWBS for 109,031 individuals aged 11-16 in Welsh Secondary schools from 2019-2021. Intercept, Estimates and their 95% confidence intervals are included for each of the 4 models (2019 survey).

| Emotional difficulties | | | | | | | | | | | | |
| --- | --- | --- | --- | --- | --- | --- | --- | --- | --- | --- | --- | --- |
|  | Model 1 | | | Model 2 | | | Model 3 | | | Model 4 | | |
|  | *b*  (95%CI) | *p* | | *b*  (95%CI) | | *p* | *b*  (95%CI) | | *p* | *b*  (95%CI) | | *p* |
| *Intercept* | 25.39  (25.33, 25.46) | <2e-16 | | 25.99  (25.91, 26.06) | | <2e-16 | 25.06  (24.91, 25.21) | | <2e-16 | 26.11  (25.96, 26.27) | | <2e-16 |
| *SDQ (0-4)*  *Close to average* | Reference group | | | Reference group | | | Reference group | | | Reference group | | |
| *SDQ (5)*  *Slightly raised* | -3.04  (-3.15, -2.93) | <2e-16 | | -2.83  (-2.94, -2.72) | | <2e-16 | -1.95  (-2.05, -1.85) | | <2e-16 | -1.58  (-1.68, -1.48) | | <2e-16 |
| *SDQ (6)*  *High* | -4.08  (-4.19, -3.98) | <2e-16 | | -3.76  (-3.87, -3.65) | | <2e-16 | -2.63  (-2.73, -2.52) | | <2e-16 | -2.14  (-2.25, -2.04) | | <2e-16 |
| *SDQ (7-10)*  *Very high* | -6.59  (-6.66, -6.51) | <2e-16 | | -6.03  (-6.12, -5.95) | | <2e-16 | -4.36  (-4.44, -4.27) | | <2e-16 | -3.45  (-3.54, -3.37) | | <2e-16 |
| Conduct problems | | | | | | | | | | | | |
|  | Model 1 | | | Model 2 | | | Model 3 | | | Model 4 | | |
|  | *b*  (95%CI) | | *p* | *b*  (95%CI) | *p* | | *b*  (95%CI) | *p* | | *b*  (95%CI) | *P* | |
| *Intercept* | 23.98  (23.91, 24.05) | | <2e-16 | 25.75  (25.67, 25.83) | <2e-16 | | 24.94  (24.79, 25.10) | <2e-16 | | 26.08  (25.93, 26.24) | <2e-16 | |
| *SDQ (0-3)*  *Close to average* | Reference group | | | Reference group | | | Reference group | | | Reference group | | |
| *SDQ (4)*  *Slightly raised* | -2.70  (-2.81, -2.59) | | <2e-16 | -2.64  (-2.75, -2.54) | <2e-16 | | -1.67  (-1.77, -1.57) | <2e-16 | | -1.01  (-1.11, -0.91) | <2e-16 | |
| *SDQ (5)*  *High* | -3.59  (-3.72, -3.47) | | <2e-16 | -3.49  (-3.61, -3.37) | <2e-16 | | -2.30  (-2.42, -2.19) | <2e-16 | | -1.41  (-1.52, -1.30) | <2e-16 | |
| *SDQ (6-10)*  *Very high* | -4.59  (-4.72, -4.46) | | <2e-16 | -4.32  (-4.44, -4.20) | <2e-16 | | -2.72  (-2.84, -2.60) | <2e-16 | | -1.56  (-1.68, -1.45) | <2e-16 | |
| Hyperactivity | | | | | | | | | | | | |
|  | Model 1 | | | Model 2 | | | Model 3 | | | Model 4 | | |
|  | *b*  (95%CI) | | *p* | *b*  (95%CI) | *P* | | *b*  (95%CI) | *p* | | *b*  (95%CI) | *p* | |
| *Intercept* | 24.69  (24.62, 24.76) | | <2e-16 | 26.03  (25.94, 26.11) | <2e-16 | | 25.07  (24.92, 25.23) | <2e-16 | | 26.20  (26.05, 26.36) | <2e-16 | |
| *SDQ (0-5)*  *Close to average* | Reference group | | | Reference group | | | Reference group | | | Reference group | | |
| *SDQ (6)*  *Slightly raised* | -2.58  (-2.69, -2.47) | | <2e-16 | -2.39  (-2.49, -2.28) | <2e-16 | | -1.47  (-1.57, -1.37) | <2e-16 | | -1.07  (-1.17, -0.98) | <2e-16 | |
| *SDQ (7)*  *High* | -3.29  (-3.40, -3.27) | | <2e-16 | -2.95  (-3.07, -2.84) | <2e-16 | | -1.83  (-1.93, -1.73) | <2e-16 | | -1.31  (-1.41, -1.22) | <2e-16 | |
| *SDQ (8-10)*  *Very high* | -5.17  (-5.26, -5.08) | | <2e-16 | -4.63  (-4.72, -4.54) | <2e-16 | | -3.02  (-3.12, -2.93) | <2e-15 | | -2.16  (-2.25, -2.06) | <2e-16 | |
| Peer problems | | | | | | | | | | | | |
|  | Model 1 | | | Model 2 | | | Model 3 | | | Model 4 | | |
|  | *b*  (95%CI) | | *p* | *b*  (95%CI) | *P* | | *b*  (95%CI) | *p* | | *b*  (95%CI) | *P* | |
| *Intercept* | 24.59  (24.52, 24.66) | | <2e-16 | 26.31  (26.22, 26.39) | <2e-16 | | 25.59  (25.44, 25.75) | <2e-16 | | 26.39  (26.24, 26.55) | <2e-16 | |
| *SDQ (0-2)*  *Close to average* | Reference group | | | Reference group | | | Reference group | | | Reference group | | |
| *SDQ (3)*  *(Slightly raised* | -2.59  (-2.68, -2.50) | | <2e-16 | -2.44  (-2.53, -2.35) | <2e-16 | | -1.65  (-1.73, -1.56) | <2e-16 | | -1.20  (-1.28, -1.12) | <2e-16 | |
| *SDQ (4)*  *High* | -3.94  (-4.05, -3.83) | | <2e-16 | -3.73  (-3.84, -3.63) | <2e-16 | | -2.61  (-2.71, -2.51) | <2e-16 | | -1.77  (-1.87, -1.67) | <2e-16 | |
| *SDQ (5-10)*  *Very high* | -5.48  (-5.58, -5.37) | | <2e-16 | -5.19  (-5.29, -5.08) | <2e-16 | | -3.72  (-3.81, -3.62) | <2e-16 | | -2.43  (-2.53, -2.32) | <2e-16 | |
| Prosocial behaviour | | | | | | | | | | | | |
|  | Model 1 | | | Model 2 | | | Model 3 | | | Model 4 | | |
|  | *b*  (95%CI) | | *p* | *b*  (95%CI) | *P* | | *b*  (95%CI) | *P* | | *b*  (95%CI) | *P* | |
| *Intercept* | 23.70  (23.63, 23.78) | | <2e-16 | 25.71  (25.62, 25.79) | <2e-16 | | 25.36  (25.20, 25.52) | <2e-16 | | 26.37  (26.21, 26.53) | <2e-16 | |
| *SDQ (7-10)*  *Close to average* | Reference group | | | Reference group | | | Reference group | | | Reference group | | |
| *SDQ (6)*  *Slightly raised* | -1.32  (-1.42, -1.21) | | <2e-16 | -1.61  (-1.71, -1.51) | <2e-16 | | -1.21  (-1.30, -1.12) | <2e-16 | | -0.80  (-0.89, -0.71) | <2e-16 | |
| *SDQ (5)*  *High* | -1.90  (-2.02, -1.78) | | <2e-16 | -2.23  (-2.35, -2.12) | <2e-16 | | -1.63  (-1.73, -1.52) | <2e-16 | | -1.00  (-1.11, -0.90) | <2e-16 | |
| *SDQ (0-4)*  *Very high* | -3.01  (-3.12, -2.90) | | <2e-16 | -3.40  (-3.50, -3.29) | <2e-16 | | -2.43  (-2.54, -2.33) | <2e-16 | | -1.41  (-1.51, -1.31) | <2e-16 | |

Note: Model 1: unadjusted model; Model 2: adjusted for demographic variables; Model 3: further adjusted for behavioural variables; Model 4: further adjusted for social variables. P values are adjusted for Bonferroni correction.
